# Optimizing training programs for athletic performance: a Monte-Carlo Tree Search variant method

**DOI:** 10.1101/2020.10.31.20223768

**Authors:** Nicolas Houy

## Abstract

**Purpose:** Using a variant of the Monte-Carlo Tree Search (MCTS) algorithm, we compute optimal personalized and generic training programs for athletic performance.

**Methods:** We use a non-linear performance model with population variability for athletes and non-athletes previously used in the literature. Then, we simulate an *in-silico* test population. For each individual of this population, we compute the performance obtained after implementing several widely used training programs as well as the one obtained by our variant of the MCTS algorithm. Two cases are considered depending on individual parameters being observed and personalized programs being possible or only parameter distributions being available and only generic training programs being implementable.

**Results:** Compared to widely used training programs, our optimization leads to an increase in performance between 1.1 (95% CI: 0.9 – 1.4) percentage point of the performance obtained with stationary optimal training dose (pp POTD) for athletes and unknown individual characteristics to 10.0 (95% CI: 9.6 – 10.3) pp POTD for nonathletes and known individual characteristics. The value of information when using MCTS optimized training strategies, *i.e*. the difference between the performance that can be reached with knowledge of individual characteristics and the performance that can be reached without it is 14.7 (95% CI: 12.8 – 16.7) pp POTD for athletes and 3.0 (95% CI: 2.6 – 3.4) pp POTD for non-athletes.

## 1 Introduction

Exercise and sport prescription is fundamental for health and fitness but also disease or injury recovery as well as professional training. For all these uses, a subject has a given objective and should plan her practice accordingly. In order to help subjects optimizing their training strategy, mathematical models have been developed. They help better understand the link between training and performance and in doing so allow a better course of action. One of these models is the Banister Impulse-Response (BIR) Model or Fitness-Fatigue Model [1, 9, 10, 13]. In this mainly phenomenological model,^1^ training implies fitness but also fatigue and both should be managed together for an optimal performance. Even though the BIR model provides a coherent framework for sport exercise studies and hence provides a useful tool for training planning and optimization, it has been also criticized for its lack of predictive ability [6, 8, 11, 26]. One of the main changes proposed in the literature since the seminal studies is the introduction of non-linearities in the way performance (through either of or both fitness and fatigue terms) reacts to different training loads [4, 5, 12, 24, 31].

This way, the BIR model becomes more realistic but optimizing its output is generally more complex. As a consequence, optimization in this field of research has been restricted to predefined sets of training programs or training strategies. Among these strategies, the ones that have been most studied without a doubt are the training strategies with tapering training loads before a competition date [10, 19–25, 27–31]. This tapering period allows to decrease both fatigue and fitness but with a profitable balance if done for the right amount of time. Less known, yet consistently described in the literature is the advantage of a work overload period before the tapering period [10, 20, 23, 27–29, 31]. Finally, even less known, a third qualitative advantageous prescription is a slight workload increase at the end of the tapering period [29, 31].

Our contribution in the present article is mainly methodological and two-fold. Using a non-linear variant of the BIR model that has been introduced before, we theoretically optimize the training programs of subjects with the objective of the highest possible performance at a given date. In order to do this, first, we insist on the degree of information available. In the first informational context, the training program designer does not have access to personal parameter values for the subjects but only population distributions. This case can occur when a prescription must be made before personalized tests have been run by the subject. It can be useful for a general web application or in any “one size fits all” context. In the second informational context, the training prescription maker has access to individual characteristics and can issue personalized prescriptions. By comparing the maximum performances obtained in both treatments – informed and non-informed about individual parameters –, we have access to the value of information, *i.e*. the increase of performance that can be obtained by a better knowledge of each subject. To the best of our knowledge, this value of information has never been quantified and yet is of importance since acquiring knowledge has a cost (sum of monetary costs for an accurate measure, costs of invasive interventions in the case of physiological measures, organizational costs for physical tests, costs for a delayed intervention, cost of giving in personal information for the patient, and more…) and this cost should certainly be compared to the corresponding expected value.

The second interest of our study is the introduction of a variant of the Monte-Carlo Tree Search (MCTS) algorithm [3] in order to find optimal training programs. The MCTS algorithm is widely used in artificial intelligence and the variant we use here has been recently implemented and proved useful in different contexts: the use of drugs to mitigate the spread of an infecting organism and the emergence and spread of resistance [14], drug regimen design in oncology [15, 18], immunotherapy [16] and chemotherapy [17].

We show, using different parameter values previously cited in the literature for different sport champions or non-athletes, that the use of our optimization algorithm can improve performance by 1.1 (95% CI: 0.9 – 1.4) pp POTD^2^ (for athletes) to 9.7 (95% CI: 9.4 – 10.0) pp POTD (for non-athletes) in the context of no knowledge of subject characteristics and by 5.1 (95% CI: 4.7 – 5.5) pp POTD (for athletes) to 10.0 (95% CI: 9.6 – 10.0) pp POTD (for non-athletes) when personalized training programs are designed. Qualitatively, the optimized training programs we issue all show (i) a tapering workload period (ii) preceded by training overload and (iii) a slight increase of the workload right before the target date, which is the date at which the performance should be as high as possible in our objective function. Hence, even though our algorithm is totally *a priori* free and has an agnostic approach regarding exercise prescription, we qualitatively observe what is usually intuitively prescribed. Moreover, with our algorithm, the quantitative aspects (when should the workload tapering period start, what shape should this tapering have, when and by how much should workload increase again…) are better founded and more precisely characterized for a better performance.

Finally, we show, that the value of information, *i.e*., again, the value of knowing the individual parameters rather than population distribution and hence, being able to design training programs accordingly, is a performance gain of 14.7 (95% CI: 12.8 – 16.7) pp POTD for athletes and 3.0 (95% CI: 2.6 – 3.4) pp POTD for non-athletes. If compared with the figures given above, information has a large value for athletes (for a point of comparison, its value is larger than using MCTS algorithm for performance optimizing) but much less for non-athletes (its value is much smaller than using MCTS algorithm for performance optimizing).

## 2 Model

### 2.1 Performance model

The performance model we build on is the non-linear variant of the Banister Impulse-Response model [1, 9] introduced in [4] and widely used since. In this model, performance at a given date *n* is the sum of a basic level *p*^0^ – obtained without ever training – and the response to training impulses (*w*_*j*_)_*j*∈_ ⟦_0,*n*−1_⟧ where *w*_*j*_ is the training load at date *j*. Formally,

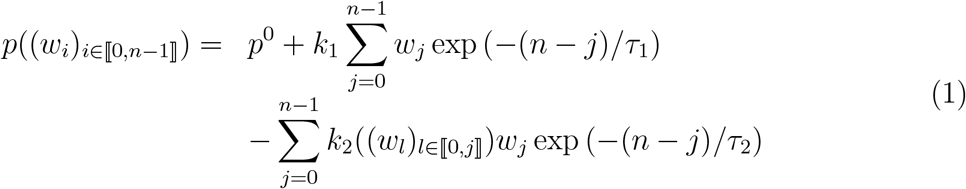

with the *k*_2_ function

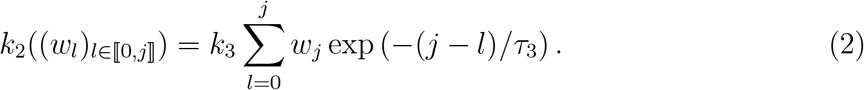

Then, response to training is the sum of a positive term (fitness) and a negative one (fatigue), both of which have a magnitude proportionately increasing with the amount of training and exponentially decreasing with time between the training date and the measurement date. Non-linearity exists because the marginal increase in fatigue is itself increasing with training loads (*k*_2_ increasing, see Equation 2).

For the sake of conciseness, let us introduce the following notation. ∀*h* ∈ [1, 3], *T*_*h*_ = exp (−1/*τ*_h_) and 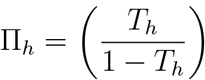.

It is straightforward to show that the performance obtained for a constant training dose *w* repeated infinitely is given by^3^

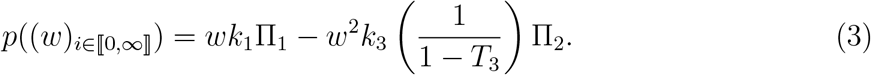

This constant training dose performance reaches its maximum value for a training dose –the optimal training dose – that we denote *OTD*

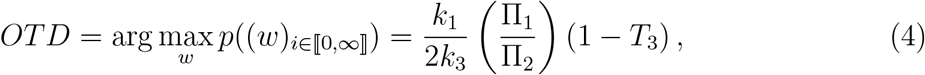

and we denote this performance value – the performance at the optimal training dose – *POTD*

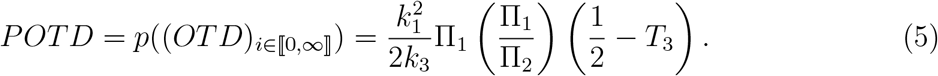

Notice that a constant training amount above 2.*OTD* yields a negative performance as fatigue becomes permanently larger than fitness. We will denote *OTD*^−^ = 2.*OTD* this training amount, above which the fatigue effect is stronger that the fitness one at stationary state.

The performance model depends on 5 parameters: *k*_1_, *k*_3_ the gain term for the initial increase in fitness and the gain term for the marginal effect of training on fatigue respectively and *τ*_1_, *τ*_2_, *τ*_3_, the decay time constants of fitness, fatigue and the marginal effect of training on fatigue respectively. In the current work, we will use the population parameter values distributions given in [4, 27, 29] for athletes and non-athletes. These parameter values are displayed in Table 1.^4^

**Table 1:** Parameter distributions for populations of athletes and non-athletes.

| Parameter | Athletes | Non-athletes |
| --- | --- | --- |
| $k_1$ | $\mathcal{N}(3.42 \times 10^{-2}, 9.6 \times 10^{-3})$ | $\mathcal{N}(3.1 \times 10^{-2}, 7.3 \times 10^{-3})$ |
| $k_3$ | $\mathcal{N}(3.4 \times 10^{-3}, 3.3 \times 10^{-3})$ | $\mathcal{N}(3.5 \times 10^{-5}, 10^{-5})$ |
| $\tau_1$ | $\mathcal{N}(43.6, 12.1)$ | $\mathcal{N}(30.8, 1.6)$ |
| $\tau_2$ | $\mathcal{N}(12.7, 7.2)$ | $\mathcal{N}(16.8, 3.3)$ |
| $\tau_3$ | $\mathcal{N}(4.1, 1.6)$ | $\mathcal{N}(2.3, 1)$ |

In order to compute different training strategies performances and compare them, we simulated two populations of 480 athletes and 480 non-athletes with parameters randomly drawn from the distributions given in Table 1. For informational purpose, OTD and POTD for these *in-silico* populations are displayed in Figure 1. The empirical average and standard deviation for athletes OTD are 18.99 and 114.62 respectively. They are 381.54 and 284.56 for non-athletes. The empirical average and standard deviation for athletes POTD are 12.81 and 66.02 respectively. They are 192.60 and 170.20 for non-athletes.

**Figure 1:**
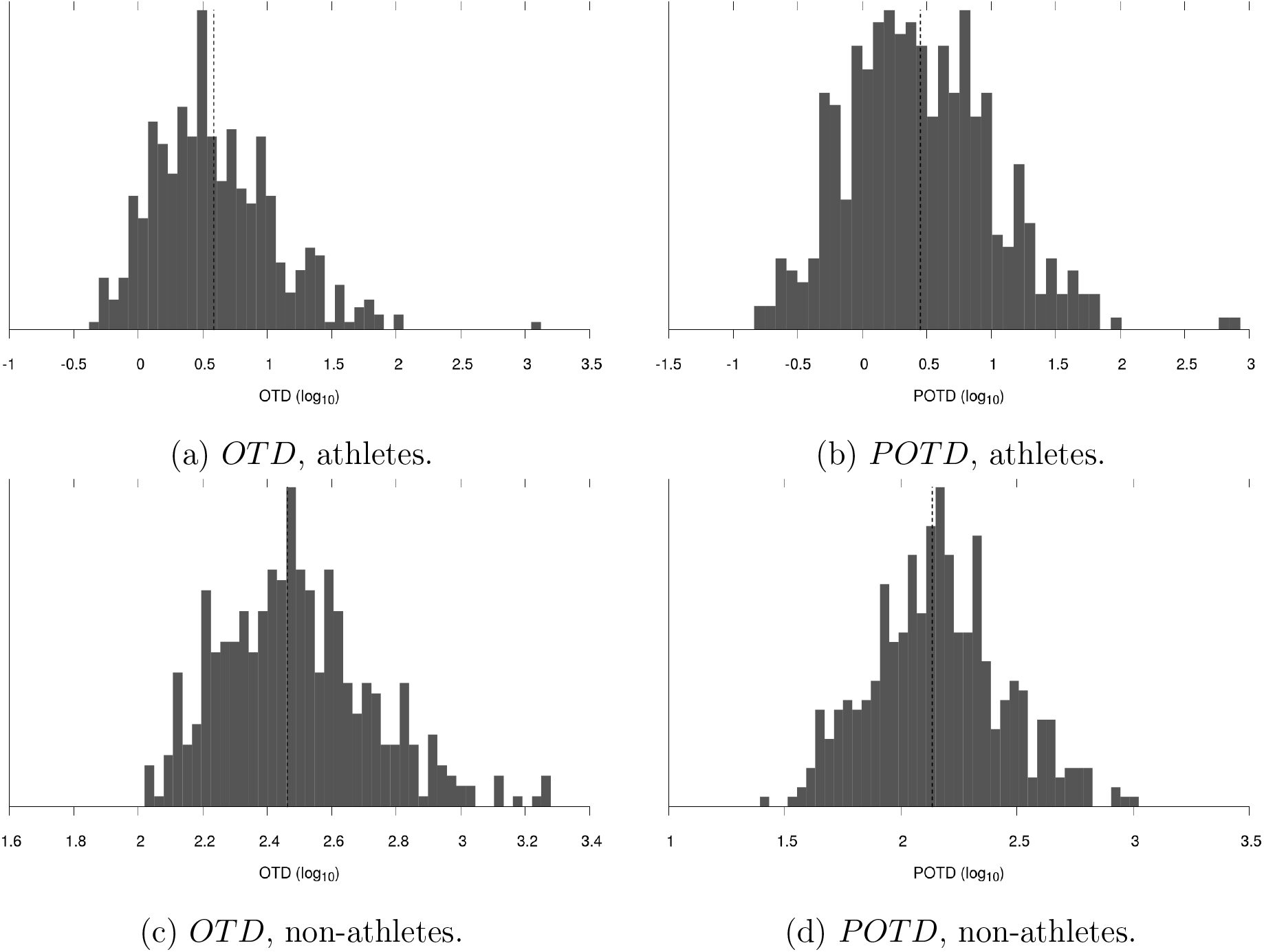
*OTD* and *POTD* empirical distributions for the 480 simulated athletes and 480 non-athletes. Dashed lines for the average individual parameter values.

Notice that for the distributions of *k*_1_ and *τ*_3_, the coefficient of variation across individuals are comparable for athletes and non-athletes. On the contrary, for the distributions of *k*_3_, *τ*_2_ and *τ*_3_, the coefficients of variation are much larger for athletes than for non-athletes. This also translates into much larger coefficients of variation for athletes than non-athletes for the distributions of *OTD* and *POTD*. Summing up roughly, the variability of athletes is much larger than the variability of non-athletes.

### 2.2 Training programs and strategies

In order to estimate the value of our optimization algorithm, we will compare it to the simple and natural training strategies that have been extensively studied in the literature and that we sum-up in the following. Notice that this list is certainly not exhaustive, yet we chose to keep its elements based on simplicity, performance and representativeness arguments.

#### OTDshare

The OTDshare(*q,n*) training program consists of a daily training with intensity *q.OTD* between day 0 and day *n* − 1, *i.e*. for *n* days.^5^

#### Step

The Step(*q, n′, n*) (with *n′* < *n*) training program consists of a daily training with intensity *q.OTD* between day 0 and day *n′* followed, if *n* − *n′* − 1 > 0,^6^ by *n* − *n′* − 1 (between days *n′* + 1 and *n* − 1) days without training or, equivalently, with intensity 0.

#### Linear

The Linear(*q, n′, n*) (with *n′* < *n*) training program consists of a daily training with intensity *q.OTD* between day 0 and day *n′* followed, if *n* − *n′* − 1 > 0,^7^ by *n* − *n′* − 1 days with training intensity linearly decreasing between *q.OTD* and 0. Formally, at day *i* ∈ [*n′* + 1, *n*− 1], the training intensity is 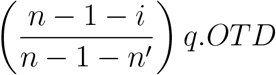.

#### Exponential

The Exponential(*q, n′, n*) (with *n′* < *n*) training program consists of a daily training with intensity *q.OTD* between day 0 and day *n′* followed, if *n* − *n′* − 1 > 0,^8^ by *n* − *n′* − 1 days with training intensity exponentially decreasing between *q.OTD* and 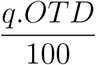. Formally, at day *i ∈* ⟦*n′* + 1, *n* − 1⟧, the training intensity is 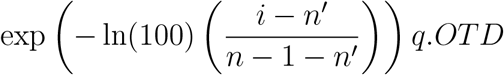.

#### MCTS

The MCTS(*q,n*) training program is the output of the optimization algorithm described in Algorithm 1.^9^ Informally, we build the MCTS training program as follows. First, we consider the set of training programs that consist on a first day (date 0) with variable training amounts and then *n* − 1 days with a default training amount, *q.OTD*. For date 0, we keep the training amount that yields the best performance at date *n* in this context. For date 1, we consider the set of training programs with the training dose found earlier at date 0, variable training amounts for date 1 and then *n* − 2 days with the default training amount *q.OTD*. For date 1, we keep the training dose that yields the best performance at date *n* in this context. We go on implementing the same procedure for *n* days. Notice that depending on the default training dose, the algorithm may yield different results.

##### Algorithm 1

Pseudo algorithm returning the MCTS training program computed for an individual with parameters *p*, a default training dose q:*OTD* and a target date n.

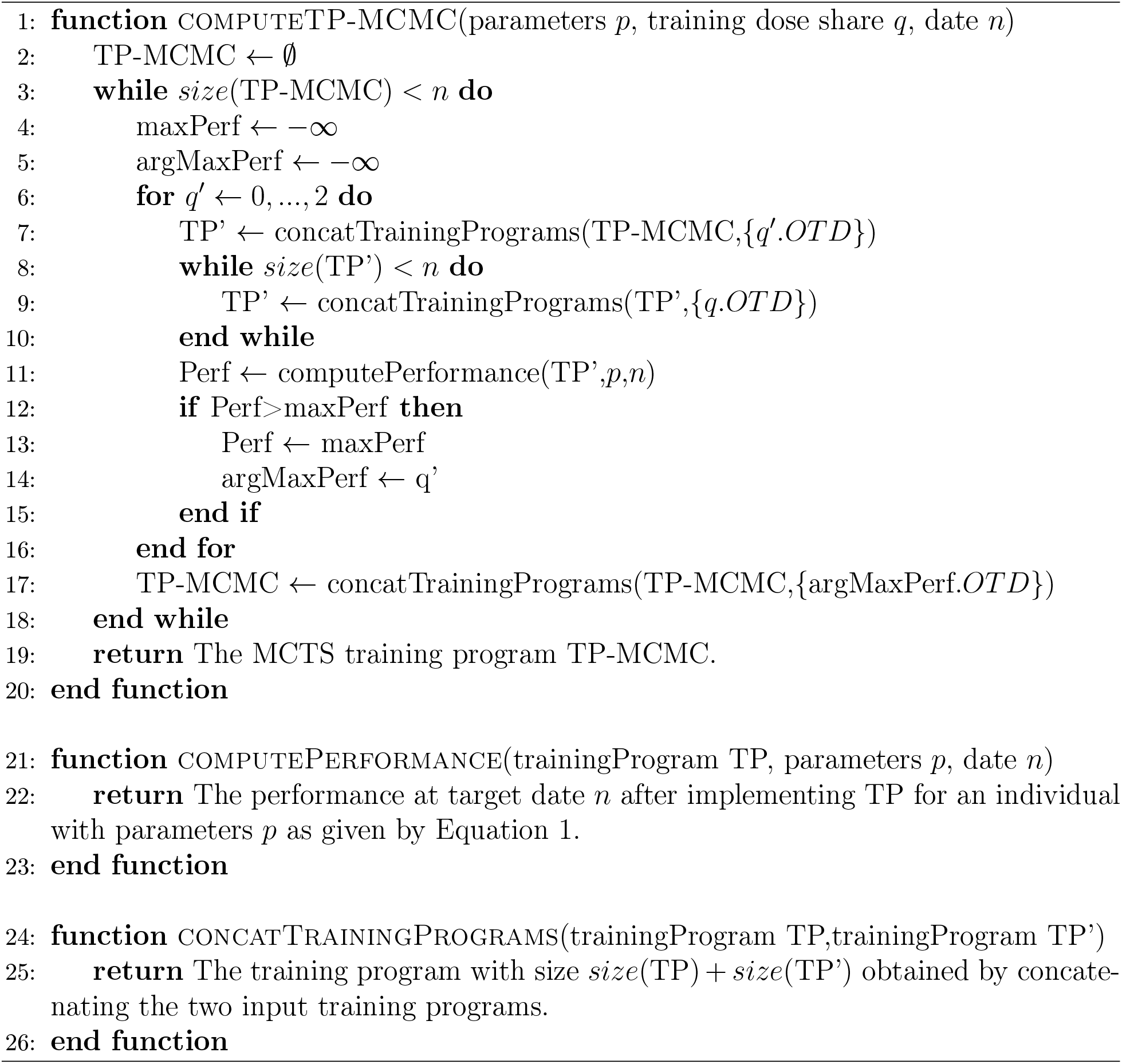

For all training programs we impose *q* between 0 and 2 which implies maximum training load of *OTD*^−^, the value above which training as a negative effect at stationary state. We call *n*, the target date which is the date at which we will compare the performances of the different training programs.

### 2.3 Information

The role of information is crucial in our study. Individual parameter values may be known for each individual or alternatively, only the parameter distributions across individuals may be known. In order to illustrate this point and its consequences, let us see how the information available to the training program designer is of importance by focusing on the OTDshare training strategy for now.

First, let us only consider an average individual. In this case, the performances of the training programs OTDshare(*q,n*) with *q* – the training dose (as a share of OTD) – and *n* –the target date – variable are displayed in Figures 2a and 2c for athletes and non-athletes respectively.^10^ From this figure, we can extract, for any target date *n*, the share of *OTD* for which the performance of the OTDshare training strategy is the best. This maximum is displayed as a plain black line in Figures 2a and 2c and the share of *OTD* which yields this maximum performance is displayed as a dashed line in Figures 3a and 3b for athletes and non-athletes respectively.

**Figure 2:**
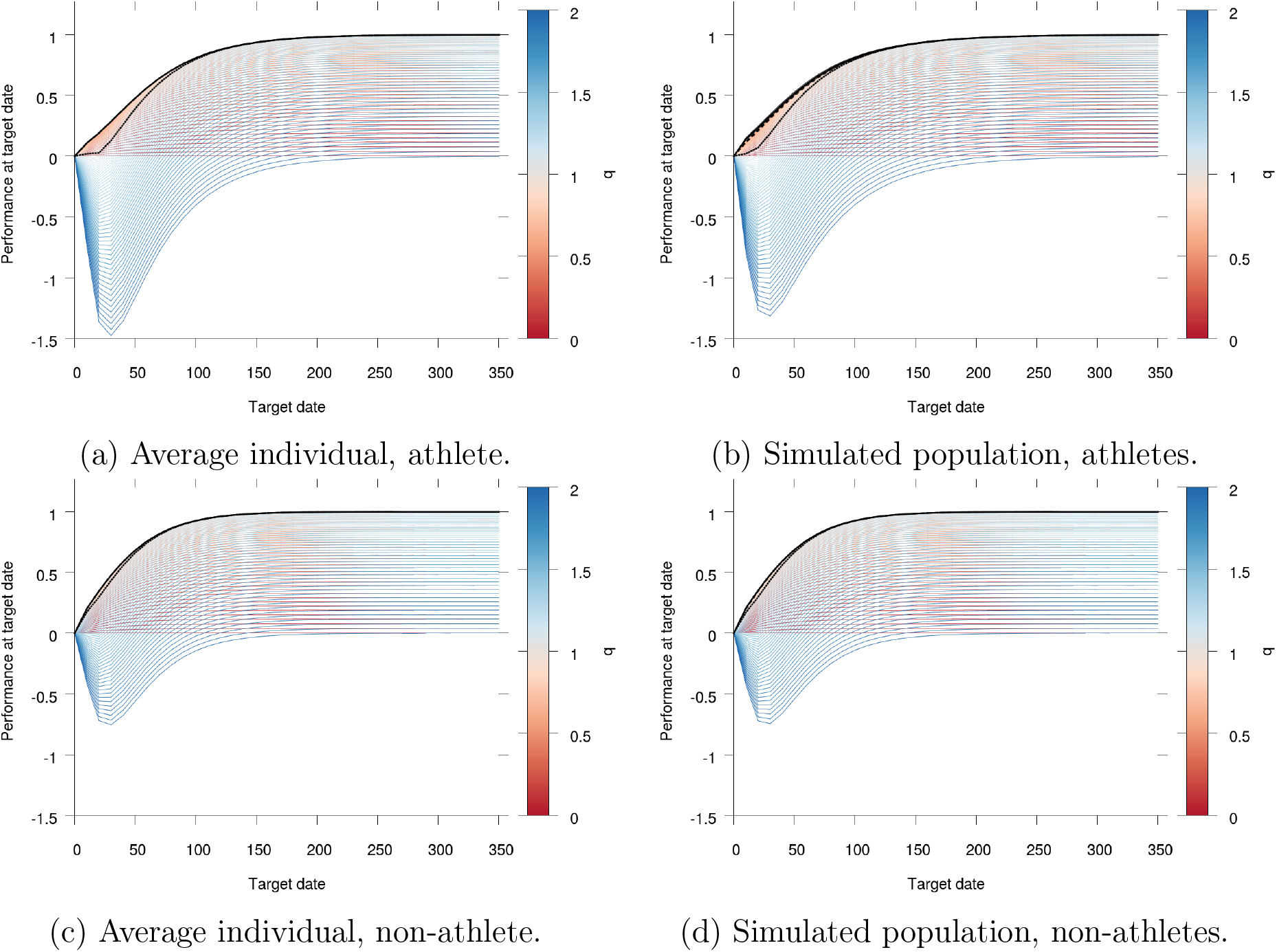
Performance of the OTDshare training strategy as a function of the target date *n* for the average athlete or non-athlete or for our simulated populations of 480 athletes and non-athletes. Color lines indicate the expected performance after completion of an OTDshare(*q,n*) training program with *q* indicated by the color of the line. The dotted line shows the expected performance of OTDshare(1,*n*). The dashed black line shows the expected performance of the best OTDshare training program when the individual parameter values are unknown, *i.e*. it shows, for each target date, the expected performance of the OTDshare training program that yields the best result for the population on average. The plain black line shows the expected performance of the best OTDshare training program when the individual parameter values are known, *i.e*. it shows, for each target date, the average performance of the best OTDshare training programs, possibly different for each individual. Notice that, by definition, the dashed and plain lines are identical when only an average individual is considered.

**Figure 3:**
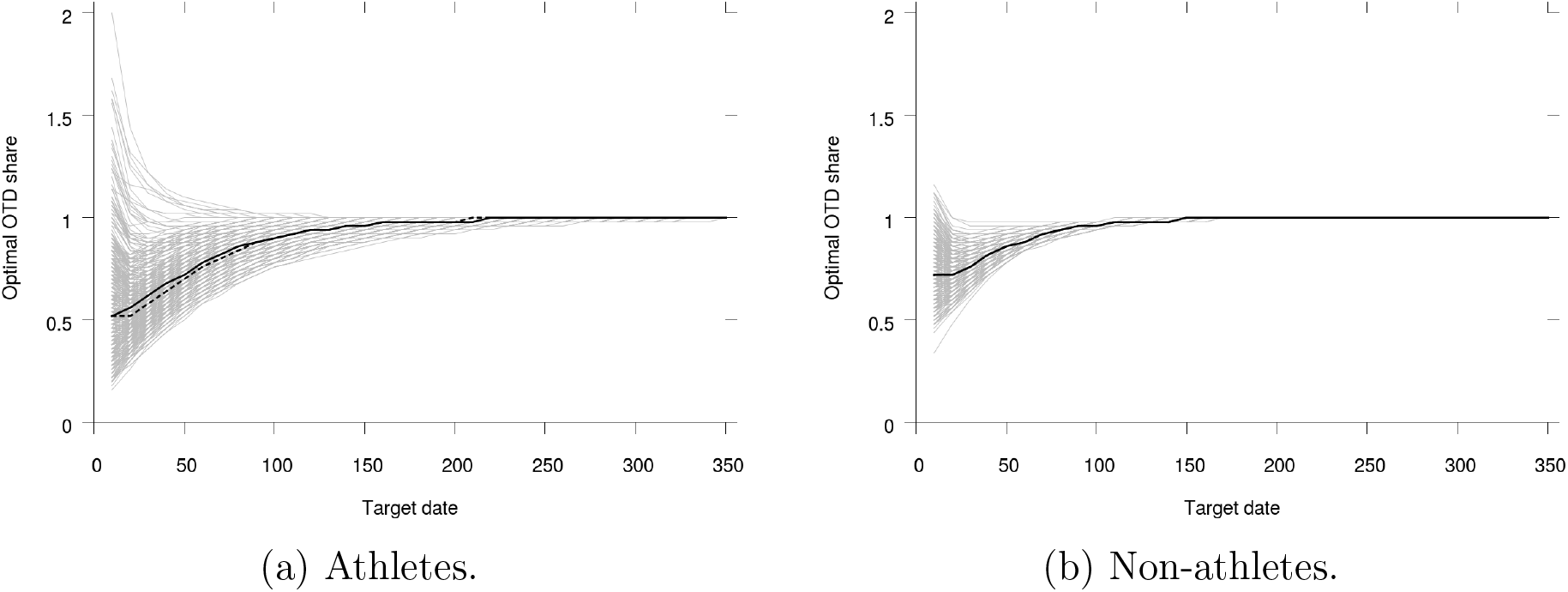
Share of *OTD* for the optimal OTDshare training programs as a function of the target date *n* for athletes and non-athletes. Light lines for each of the 480 simulated individuals in the populations. Plain black line for the optimal OTDshare training program for the populations without knowledge of individual parameter values. Dashed black line for the optimal OTDshare training program for the average individual.

Now, when we face not only the average individual but a whole population, the question arises of the knowledge of each individual parameter values. First, let us assume that the parameters are not observed or observable. Only the parameters distributions are available to the training program designer. In this case, our purpose will be to design a training program that yields the best results *on average* over the population. In Figures 2b and 2d, we show the average or expected performance of the training programs OTDshare(*q,n*) with *q* and *n* variable for our simulated populations of athletes and non-athletes respectively. In dashed lines, for each target date *n*, we display the performance of the best program of the OTDshare training strategy in this informational context. Since the individual parameters are not known, it is the training program that yields the best performance on average for a population with parameters distributed as described in Table 1. The share of *OTD* that yields this best performance is displayed in plain black line in Figures 3a and 3b for athletes and non-athletes respectively.

Now, let us assume that we know individual parameters. In this case, it may be more profitable to design a personalized training program for each individual. For the populations of athletes and non-athletes we simulated, the optimal shares of *OTD* for the OTDshare training strategy yielding the best performances (one optimal share of *OTD* for each individual) are displayed in light lines in Figures 3a and 3b. Once these personalized training programs are implemented, the expected performance for the population is given in plain black lines in Figures 2b and 2d.^11^

Notice that, by definition of *OTD*, when the target date is infinite, the best program of the OTDshare strategy is obtained for a constant training amount *OTD, i.e*. an optimal OTD share of 1. This remark holds whatever the informational context, each individual, known or not, average or part of a population, reaches at most performance *POTD* when the target date goes to infinity.

The value of information is the difference of the performances that can be reached with or without knowledge of individual parameter values.^12^ As we remarked above, the value of information tends to 0 when the target date goes to infinity. However, even though it can hardly be seen on the figures because it is very low in the case of the OTDshare training strategy, it is not necessarily 0 for small target dates. It can be as much as 2.6 (95% CI: 2.1 – 3.1) pp POTD for athletes and 0.6 (95% CI: 0.6 – 0.7) pp POTD for non-athletes (both maximum values of information are obtained for a target date of 10 days). The values of information as functions of the target day for the OTDshare training strategy are displayed in Figure 4.

**Figure 4:**
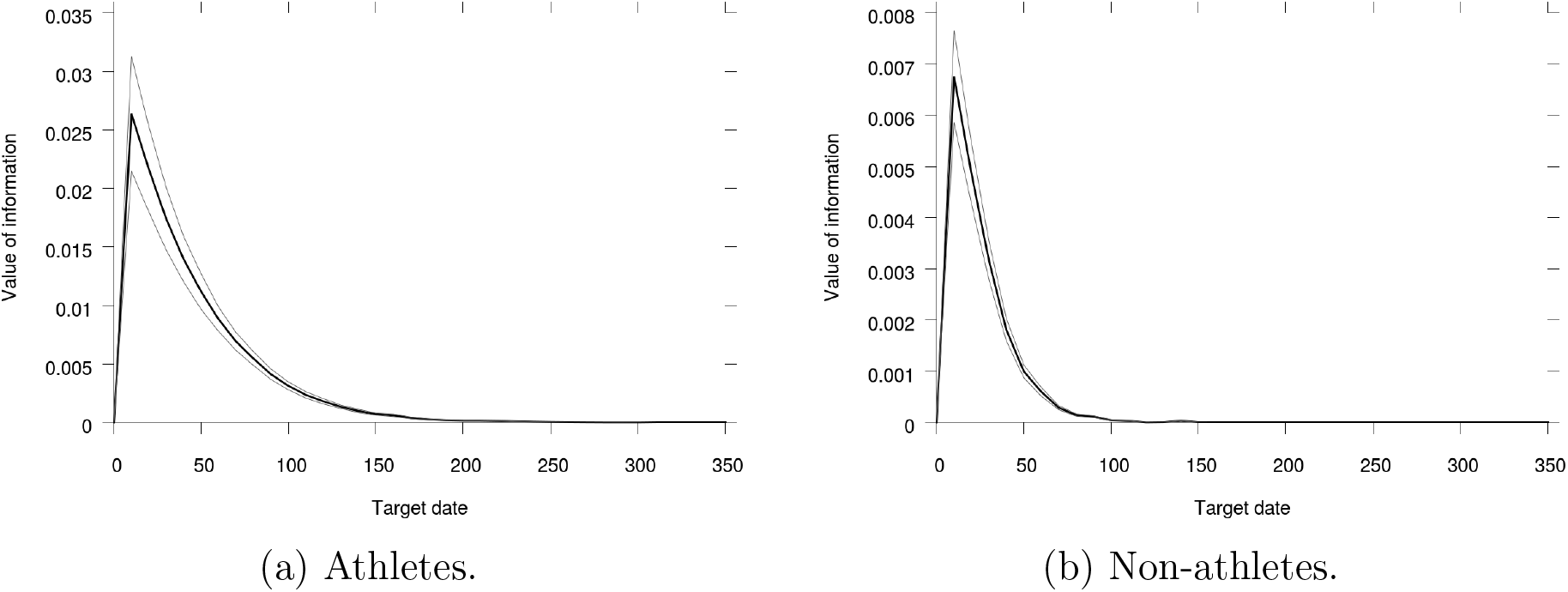
Value of information for the OTDshare training strategy as a function of the target date for the athletes and non-athletes populations. Light lines show the 95% confidence interval for our 480 simulated individuals.

Notice that because we are dealing with an *in-silico* population, when we compare two treatments (with or without knowledge of the individual parameter values in the case of Figure 4 for instance), we can compare the performances obtained by each individual with identical characteristics. Then statistics are more powerful than a real trial that would compare two populations with different parameters only.

## 3 Results

In this section, we will focus on target date 350, when the boundary effect of date 0 has vanished.^13^

### 3.1 Average individual

First, let us focus on the case with only the average individuals for both the athletes and non-athletes populations. As already noticed, at target date 350, for both the average athlete and non athlete, the best training program of the OTDshare strategy is obtained with a daily training dose of *OTD* (OTDshare(1,350)) and yields a performance of 1 (by definition of *OTD* since the stationary state is virtually reached at date 350).

For athletes, the best Step training program is Step(327,2,350) and yields a performance of 1.71, the best Linear training program is Linear(308,2,350) and yields a performance of 1.87, the best Exponential training program is Exponential(322,2,350) and yields a performance of 1.79. The best MCTS training program is obtained for a default training dose 0.52 *OTD* and it yields a performance of 1.89.

For non-athletes, the best Step training program is Step(349,1,350)^14^ and yields a performance of 1, the best Linear training program is Linear(305,2,350) and yields a performance of 1.22, the best Exponential training program is Exponential(320,2,350) and yields a performance of 1.03. The best MCTS training program is obtained for a default training dose 0.8 *OTD* and it yields a performance of 1.32.

Hence, the MCTS training strategy leads to the best performances for target date 350 in all cases.^15^ Results are summed-up as differences in performance to MCTS in Table 2.

**Table 2:** Difference in performance between the best MCTS training program and the best other training programs for an average athletes, average non-athletes and for a target date of 350.

| Training programs difference | Athletes | Non-athletes |
| --- | --- | --- |
| MCTS-OTDshare | 0.889 | 0.320 |
| MCTS-Step | 0.182 | 0.320 |
| MCTS-Linear | 0.023 | 0.103 |
| MCTS-Exponential | 0.094 | 0.288 |

The daily training doses of the best MCTS training programs for athletes and non-athletes are displayed in Figure 5. We can see that the MCTS training programs obtained by optimization qualitatively reproduce three well-known features of the literature: the work overload^16^ followed by a tapering workload period by the end of which, the training doses are increased a bit again.

**Figure 5:**
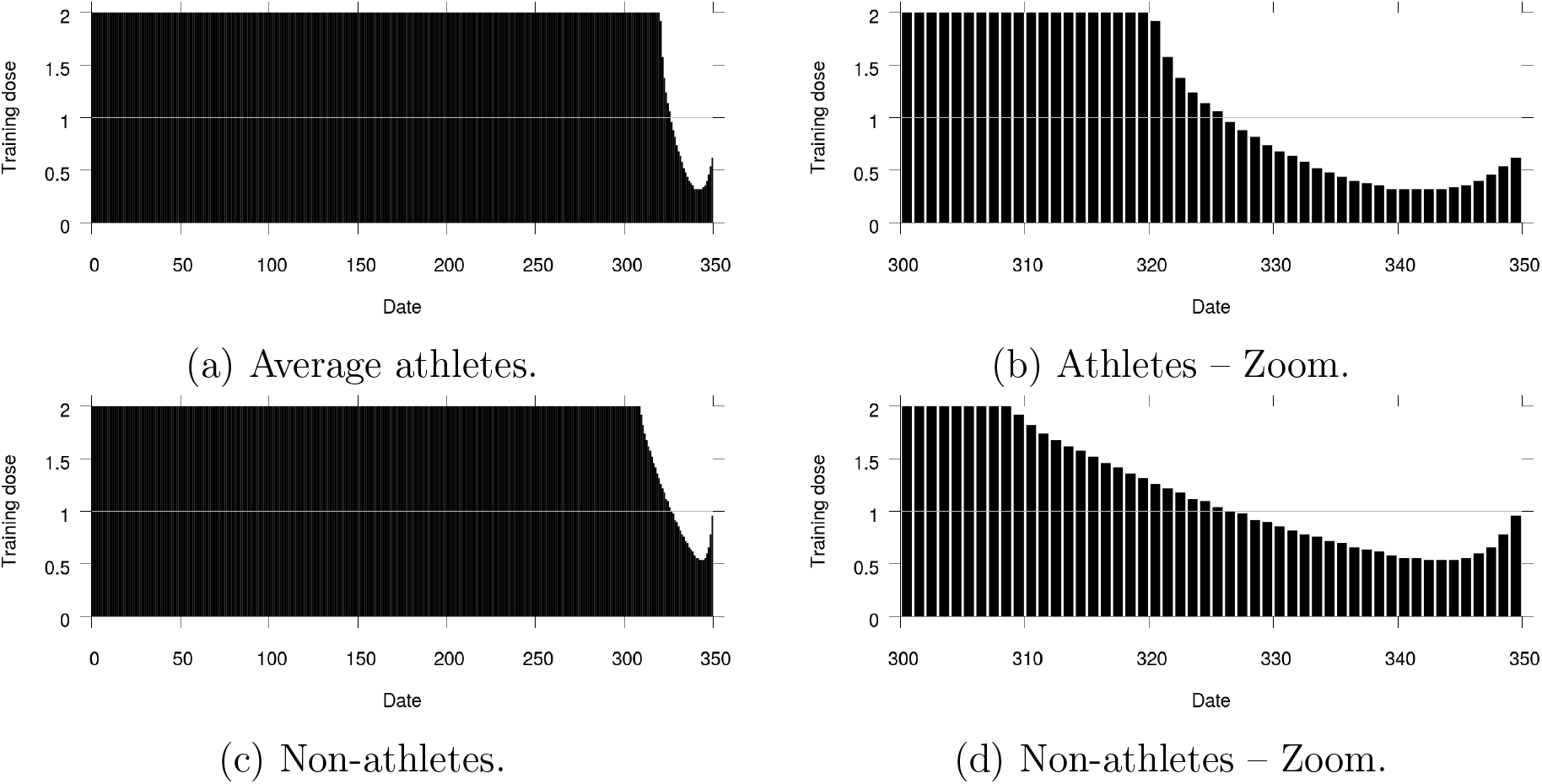
Daily training doses (*w*_*i*_)_*i*∈ ⟦0,349⟧_ obtained by the best MCTS training program for an average athlete, an average non-athlete and for target date 350.

### 3.2 Unknown individual parameter values

Let us now turn to the case where we are dealing with whole populations for which we know the parameters distributions but we cannot observe individual characteristics so that we cannot consider personalized training programs. Instead, only training programs that apply to all individuals indiscriminately can be considered and we search the ones that yield the best performances in expectation over our simulated populations.

In this case, at date 350, for both populations of athletes and non-athletes, the best OTDshare training program – *i.e*. the one that gives the highest expected performance – is obtained with a daily training dose of *OTD* (OTDshare(1,350)) and yields a performance of 1.00 (95% CI: 1.00 – 1.00), for the same reasons as above.

For athletes, the best Step training program is Step(330,2,350) and yields a performance of 1.58 (95% CI: 1.52 – 1.64), the best Linear training program is Linear(310,2,350) and yields a performance of 1.75 (95% CI: 1.71 – 1.80), the best Exponential training program is Exponential(324,2,350) and yields a performance of 1.67 (95% CI: 1.61 – 1.72). The best MCTS training program is obtained for a default training dose 0.56 *OTD* and it yields a performance of 1.77 (95% CI: 1.72 – 1.82).^17^

For non-athletes, the best Step training program is Step(349,1,350) and yields a performance of 1.00 (95% CI: 1.00 – 1.00),^18^ the best Linear training program is Linear(306,2,350) and yields a performance of 1.22 (95% CI: 1.19 – 1.24), the best Exponential training program is Exponential(322,2,350) and yields a performance of 1.04 (95% CI: 1.01 – 1.06). The best MCTS training program is obtained for a default training dose 0.88.OTD and it yields a performance of 1.31 (95% CI: 1.30 – 1.33).

Hence, the best MCTS training program is the training program that gives the best performance for target date 350. Because we are dealing with an *in-silico* population, we can compare the performances obtained by each individual when treated by each of the training strategies. The expected differences in performance between the best MCTS training program and the best programs of other strategies are given in Table 3. We can see that the gain in performances obtained using the MCTS training strategy is significant in all cases.

**Table 3:**
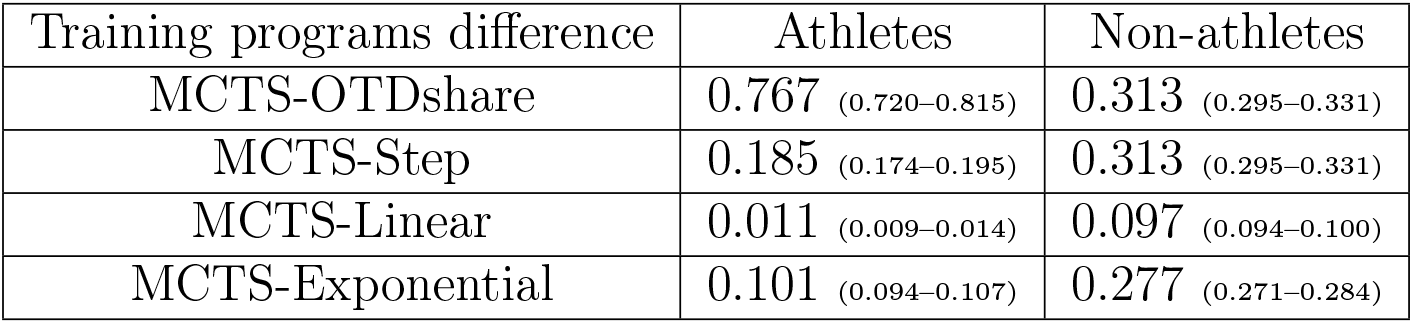
Differences in performance between the best MCTS training program and the best other training programs for athletes, non-athletes and for a target date of 350. Individual parameter values are unknown. 95% confidence interval for our 480 simulated individuals in parenthesis.

| Training programs difference | Athletes | Non-athletes |
| --- | --- | --- |
| MCTS-OTDshare | 0.767 (0.720–0.815) | 0.313 (0.295–0.331) |
| MCTS-Step | 0.185 (0.174–0.195) | 0.313 (0.295–0.331) |
| MCTS-Linear | 0.011 (0.009–0.014) | 0.097 (0.094–0.100) |
| MCTS-Exponential | 0.101 (0.094–0.107) | 0.277 (0.271–0.284) |

The best MCTS training programs for athletes and non-athletes are displayed in Figure 6. They are qualitatively very close to what was implemented for the average athlete and non-athlete. In particular, they display the overload before the tapering workload period ending with a slight increase in training dose.

**Figure 6:**
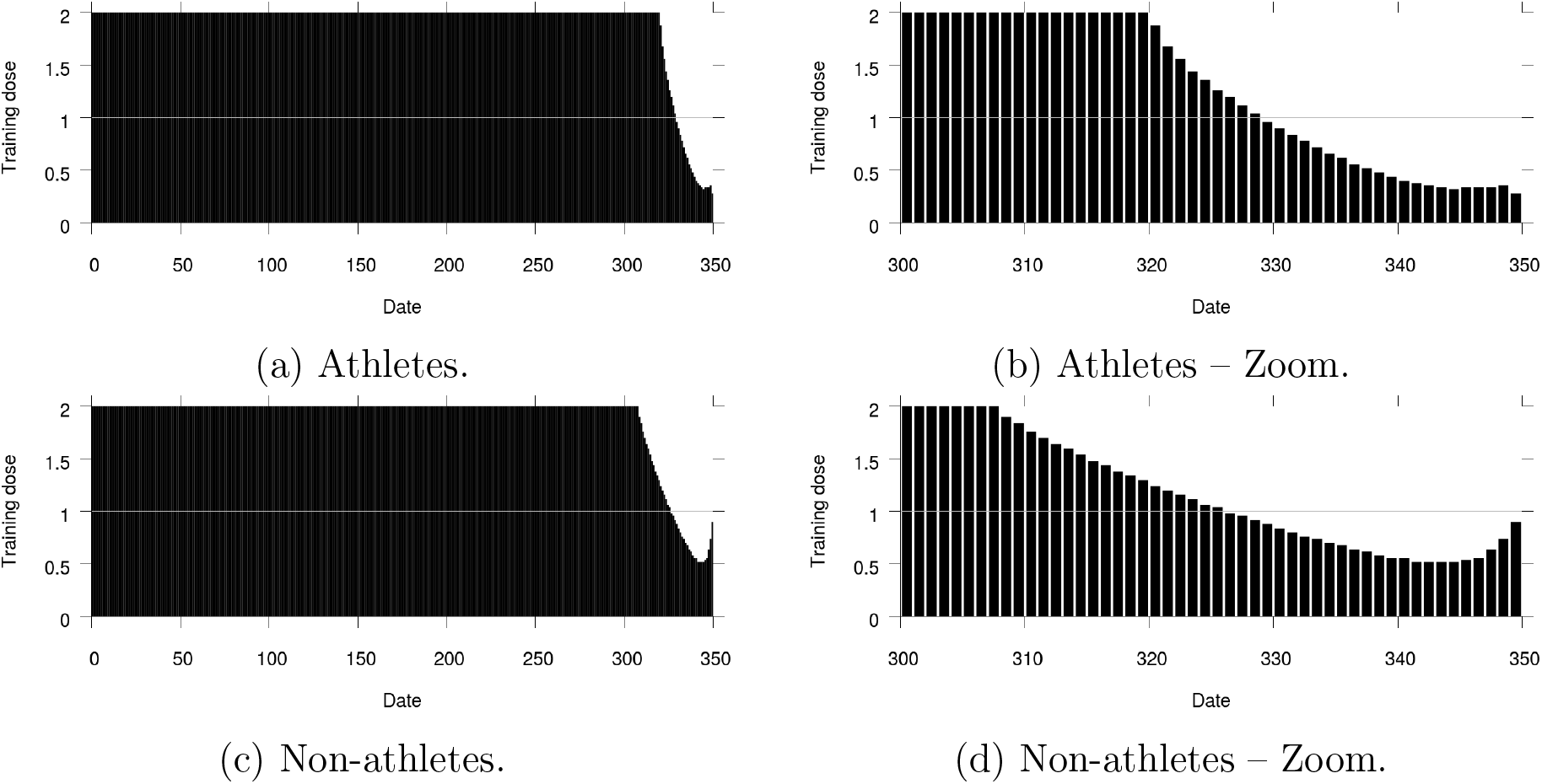
Daily training doses (*w*_*i*_)_*i*∈ ⟦0,349⟧_ obtained by the best MCTS training program for populations of athletes and non-athletes with unknown individual parameter values and for target date 350.

For informational purpose, let us consider what would be the performances obtained by our simulated populations implementing the best MCTS training programs computed in Section 3.1 – *i.e*. the training programs that were obtained when considering the average individuals only. These results are the ones that one would get if not the whole information about parameter distributions was used but only the average statistics. For athletes (respectively non-athletes), implementing the best MCTS training program computed on the average individual would yield an expected performance of 1.75 (95% CI: 1.71 – 1.80) (resp. 1.31 (95% CI: 1.30 – 1.33)). The per-individual differences in performance between the two treatments (MCTS computed on the whole population versus MCTS computed on the average individual) is 0.008 (95% CI: 0.003 – 0.014) (resp. 1.3×10^−5^ (95% CI: −9.3 ×10^−5^ – 1.2 × 10^−4^)). Hence, we can conclude that, given the populations we consider here, there is almost no cost in terms of performances to use the average individual instead of the whole population in order to compute the best MCTS training program. Yet, the gains in terms of computational power needed (and as a consequence, time of computation and/or resource usage) are large.

### 3.3 Known individual parameter values

We will now study the case where not only do we know the distributions of the parameters in the population but, we can also observe the parameter values *for each individual*. In this case, the training program to be implemented can be personalized, *i.e*. for each individual, we can reach the best performance for any training strategy.

In this case, at date 350, for both populations of athletes and non athletes and for each individual, the best OTDshare training program – *i.e*. the one that gives the highest performance – is obtained with a daily training dose of *OTD* (OTDshare(1,350)) and yields an average performance for all individuals of 1.00 (95% CI: 1.00 – 1.00). Notice this allows to check that for all individuals, date 350 is enough to avoid date 0 boundary effect.

For athletes, when her best Step training program is implemented for each individual, it yields an average performance of 1.74 (95% CI: 1.68 – 1.79), when her best Linear training program is implemented for each individual, it yields an average performance of 1.86 (95% CI: 1.81 – 1.91), when her best Exponential training program is implemented for each individual, it yields an average performance of 1.81 (95% CI: 1.75 – 1.86), when her best MCTS training program is implemented for each individual, it yields an average performance of 1.91 (95% CI: 1.86 – 1.96).

For non-athletes, when her best Step training program is implemented for each individual, it yields an average performance of 1.08 (95% CI: 1.06 – 1.09), when her best Linear training program is implemented for each individual, it yields an average performance of 1.24 (95% CI: 1.22 – 1.26), when her best Exponential training program is implemented for each individual, it yields an average performance of 1.13 (95% CI: 1.12 – 1.15), when her best MCTS training program is implemented for each individual, it yields an average performance of 1.34 (95% CI: 1.33 – 1.36).

Differences between the MCTS strategy and other strategies, when performances at date 350 are compared for each individual, are given in Table 4. Again, the gains in using MCTS are always significant.

**Table 4:** Difference in performance between the best MCTS training program and the best other training programs for athletes, non athletes and for a target date of 350. Individual parameter values are known. 95% confidence interval for our 480 simulated individuals in parenthesis.

| Training program difference | Athletes | Non-athletes |
| --- | --- | --- |
| MCTS-OTDshare | 0.915 (0.866–0.964) | 0.343 (0.326–0.360) |
| MCTS-Step | 0.175 (0.165–0.184) | 0.265 (0.257–0.273) |
| MCTS-Linear | 0.051 (0.047–0.055) | 0.100 (0.096–0.103) |
| MCTS-Exponential | 0.106 (0.099–0.113) | 0.210 (0.204–0.215) |

A representation of the best MCTS training programs for athletes and non-athletes are displayed in Figure 7. Here again, they are qualitatively very close to what was implemented for the average athlete and non-athlete and for unknown individual parameter values. Overload before a workload tapering period and a workload increase at the end of this workload tapering period can be observed.

**Figure 7:**
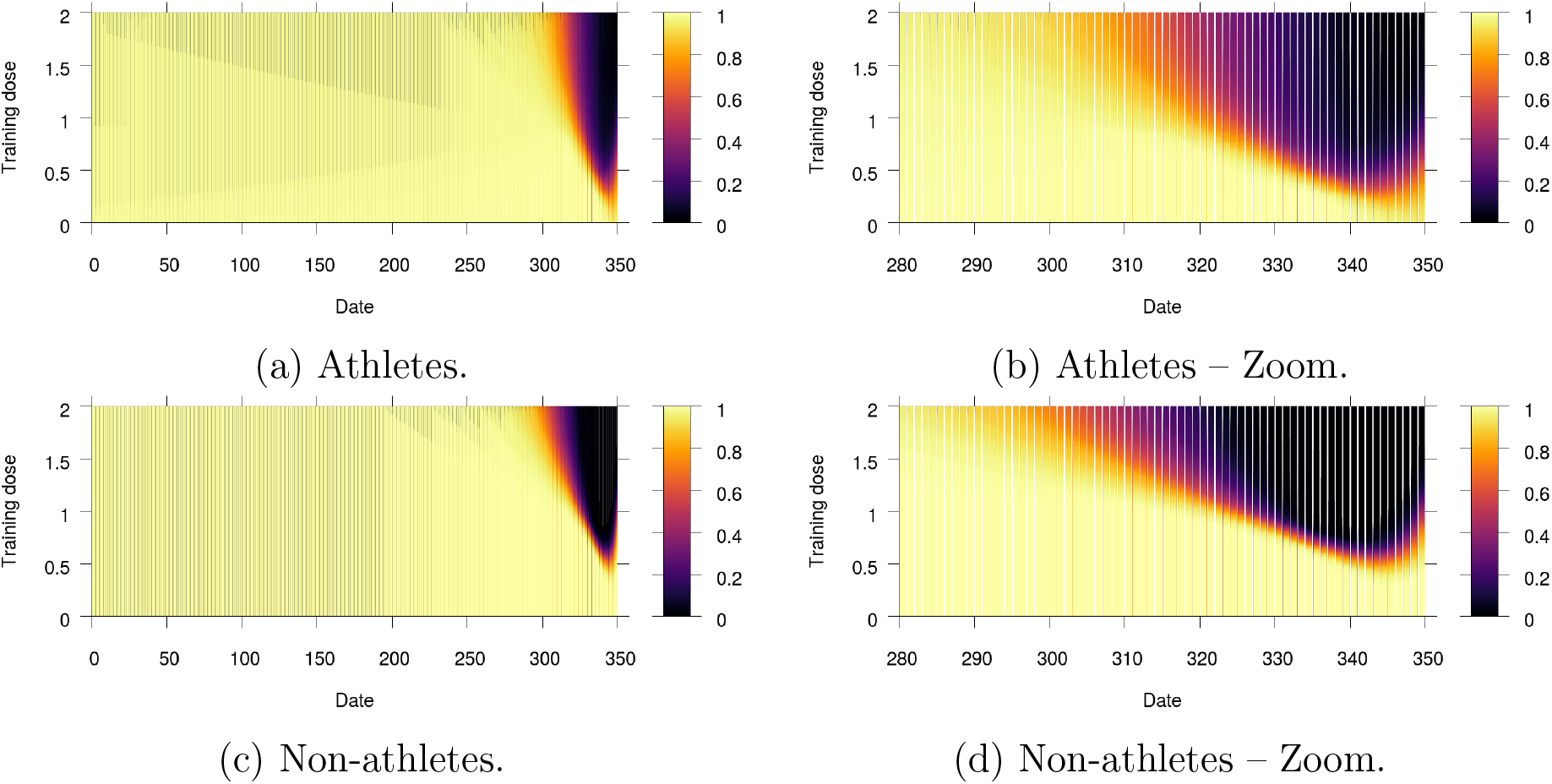
Daily training doses proportions for our simulated populations of athletes and nonathletes. For each date (abscissa), and each training dose level as a share of *OTD* (ordinate), the color indicates the proportion of individuals in our populations who are prescribed a training dose more than the ordinate training dose level at this date and when each individual implements her best MCTS training program.

### 3.4 Value of information

Finally, let us compute the value of information. As explained above, it is the value, in terms of performance gains, of having access to each individual parameter values instead of having access only to the population parameters distributions. Then, in our case, it is the difference in performance between a treatment where each individual can be trained as prescribed by her own best training program of a given strategy (as computed in Section 3.3) on the one hand and a treatment where all individuals are trained as prescribed by the same best *on average* training program of the same given strategy (as computed in Section 3.2) on the other hand.

In Section 2.3, we saw that, for the OTDshare training strategy, the value of information is quite small for all target dates and it is even tending to 0 when the target date tends to infinity by definition. These results do not hold for the other training strategies. Indeed, for the other training strategies, the value of information can be large. Knowing individual parameter values can increase the performance after training by 19.8 (95% CI: 17.7 – 21.9) pp POTD for athletes (for the Step training strategy and target date 60) and 11.3 (95% CI: 9.7 – 13.0) pp POTD for non-athletes (for the Exponential training strategy and target date 120). Also, the value of information is not necessarily tending to 0 for distant target dates. Values of information for the different strategies at target date 350 are displayed in Table 5.

**Table 5:** Value of information for different training strategies at target date 350. 95% confidence interval for our 480 simulated individuals in parenthesis.

| Training strategy | Athletes | Non-athletes |
| --- | --- | --- |
| OTDshare | 0.000 (0.000–0.000) | 0.000 (0.000–0.000) |
| Step | 0.157 (0.134–0.181) | 0.078 (0.064–0.093) |
| Linear | 0.107 (0.090–0.124) | 0.028 (0.023–0.033) |
| Exponential | 0.142 (0.120–0.164) | 0.098 (0.087–0.110) |
| MCTS | 0.147 (0.128–0.167) | 0.030 (0.026–0.034) |

We show the values of information as functions of the target date for the Step, Linear, Exponential and MCTS training strategies in Figures 8, 9, 10 and 11 respectively.

**Figure 8:**
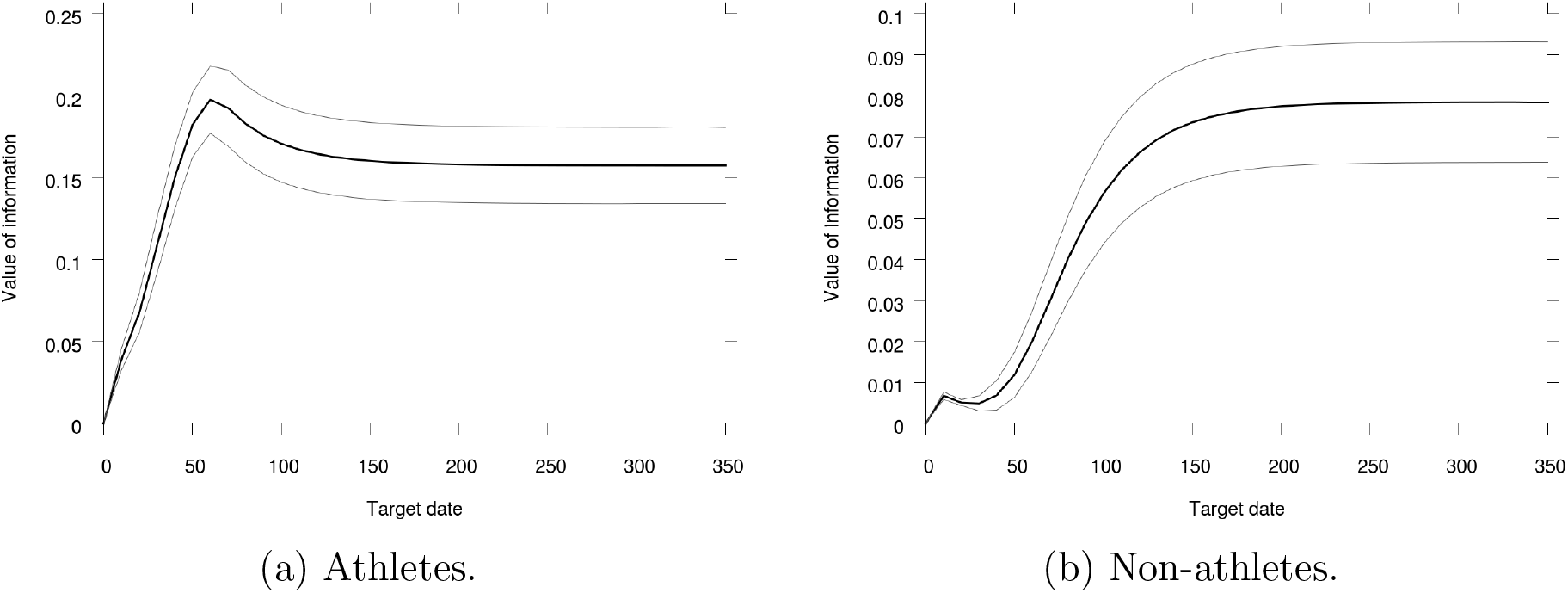
Value of information for the Step training strategy as a function of the target date for the athletes and non-athletes populations. Light lines show the 95% confidence interval for our 480 simulated individuals.

**Figure 9:**
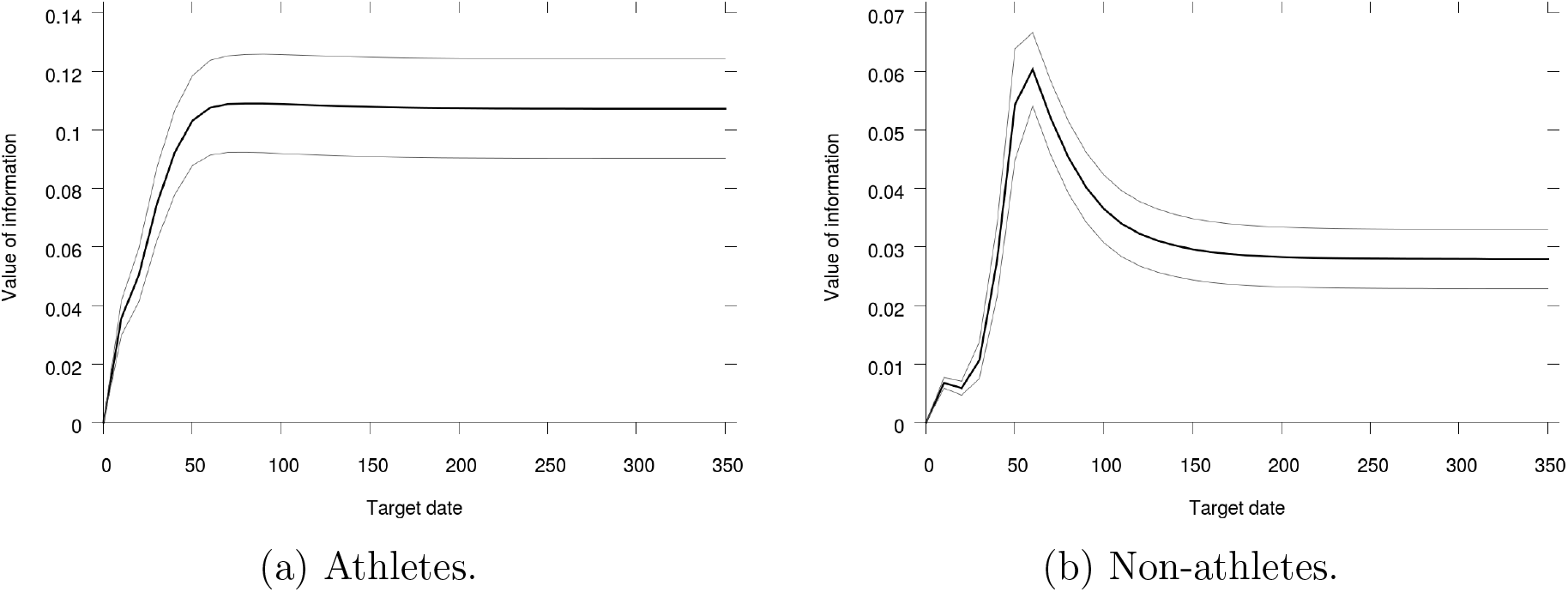
Value of information for the Linear training strategy as a function of the target date for the athletes and non-athletes populations. Light lines show the 95% confidence interval for our 480 simulated individuals.

**Figure 10:**
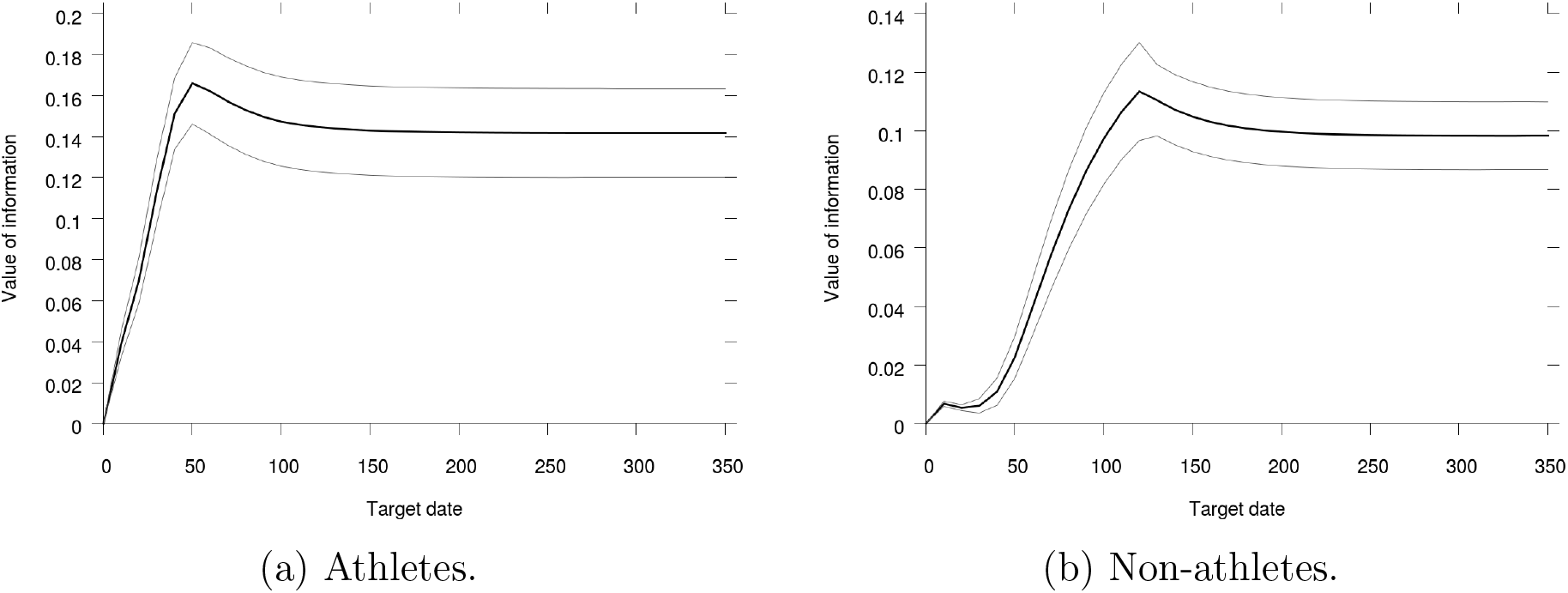
Value of information for the Exponential training strategy as a function of the target date for the athletes and non-athletes populations. Light lines show the 95% confidence interval for our 480 simulated individuals.

**Figure 11:**
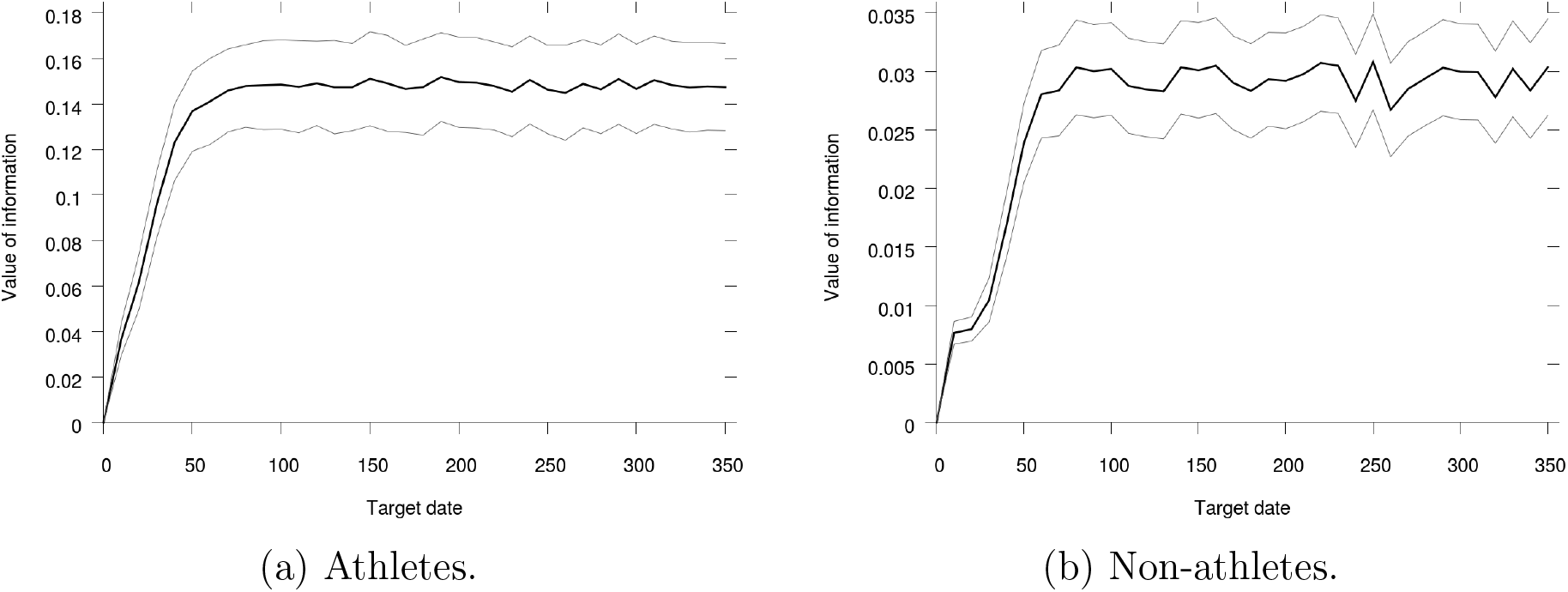
Value of information for the MCTS training strategy as a function of the target date for the athletes and non-athletes populations. Light lines show the 95% confidence interval for our 480 simulated individuals.

## 4 Discussion and conclusion

Apart from the MCTS training strategy, the best strategy for both populations of athletes and non-athletes is the Linear strategy. In Figure 12, we sum up the comparisons we found between the Linear and MCTS strategies depending on the different informational contexts we studied in this article. In this figure, we can see that a way to illustrate our results is by splitting the gains between implementing the Linear training strategy without knowledge of the individual parameter values and implementing the MCTS training strategy with knowledge of the individual parameter values into two parts: First the informational value of knowing the individual parameter values (informational component) and second, the value to use the MCTS training strategy instead of the linear one (optimization component).

**Figure 12:**
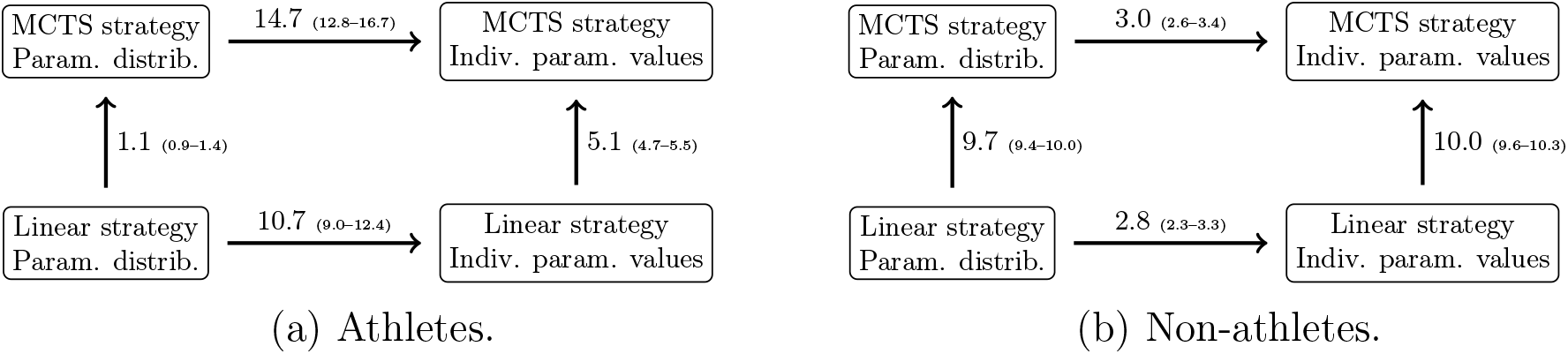
Diagram of our results for athletes and non-athletes populations. An arrow indicates an increase in performance with magnitude in pp POTD indicated as numbers close to it. 95% confidence interval for our 480 simulated individuals in parenthesis.

In the case of athletes, the performance gains come mainly from the informational component whereas in cases of non-athletes, the gains come mainly from the optimization component. The fact that the informational component is quite small in the case of the non-athletes population comes obviously from the fact that variation coefficients are quite small in this population. On the contrary, the optimization component is larger in the non-athletes population.

The optimal MCTS training programs all show (i) an overload period before a (ii) workload tapering period ending with a (iii) slight increase in training doses. Even though our results are hard to compare with previous studies using the same performance model and data [4, 27, 29] – because of the non-linearity of our tapering workload period or of the subsequent increase, the difference in the preceding overload – we qualitatively find similar results. It is to be noted that these results have been found here with an algorithm totally open to very different training strategies. Said differently, even though our algorithm had non reason to point to training programs that were previously studied in particular, we found qualitative features for optimal training programs that were already present in these previous studies. Of course our algorithm still brings quantitative precisions that could not be reached before.

Finally, we would like to highlight a feature of the MCTS training strategy that would certainly prove to be very important for future real-life implementation. The MCTS training strategies are based on Monte-Carlo simulations This implies that the MCTS training strategy is very robust to any foreseen, unforeseen event or even information acquisition or learning. For instance, let us assume that the individual for which we compute the optimal training program suffers injury. Then, it is immediate to recompute the algorithm in order to have the optimal training program being given the suffered injury and the constraints it imposes on our new problem but also the learning of injury probabilities, possibly as a function of training loads. This would not be the case for many other algorithms like a genetic one for instance [24]. Such implementations are left for future works.

## Data Availability

Nothing to report

## Appendix

### A MCTS variant for unknown individual parameter values

In Algorithm App-1, we show the variant to Algorithm 1, for a population with unknown individual parameter values. In this case, the algorithm depends only on the parameter distribution and basically, it works by first drawing by chance a population^19^ and then, implementing the 1-individual MCTS algorithm with expected performance over the drawn population as an objective function. Obviously, the population drawn in order to build the MCTS training program (training population) should not be the same as the one that was drawn in order to test the different training program treatments (validation population).

#### Algorithm App-1

Pseudo algorithm returning the MCTS training program computed for parameter distribution *d*, a default training dose *q.OTD* and a target date *n*.

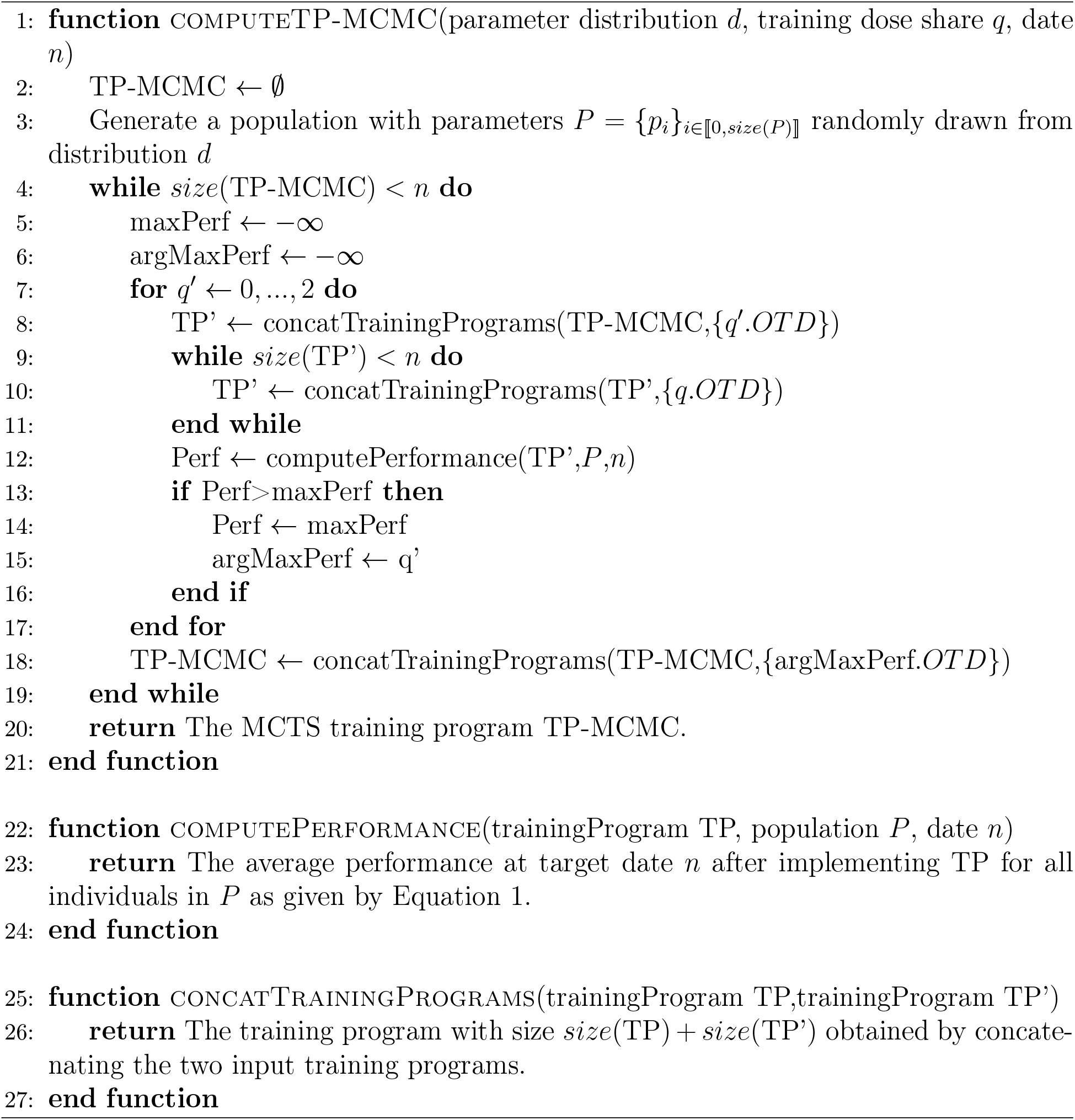

### B Best performances of the training strategies

#### B.1 Performances

In Figures App-1, App-2, App-3, App-4 and App-5, we show how the training strategies studied in this article perform depending on the target date.

**Figure App-1:**
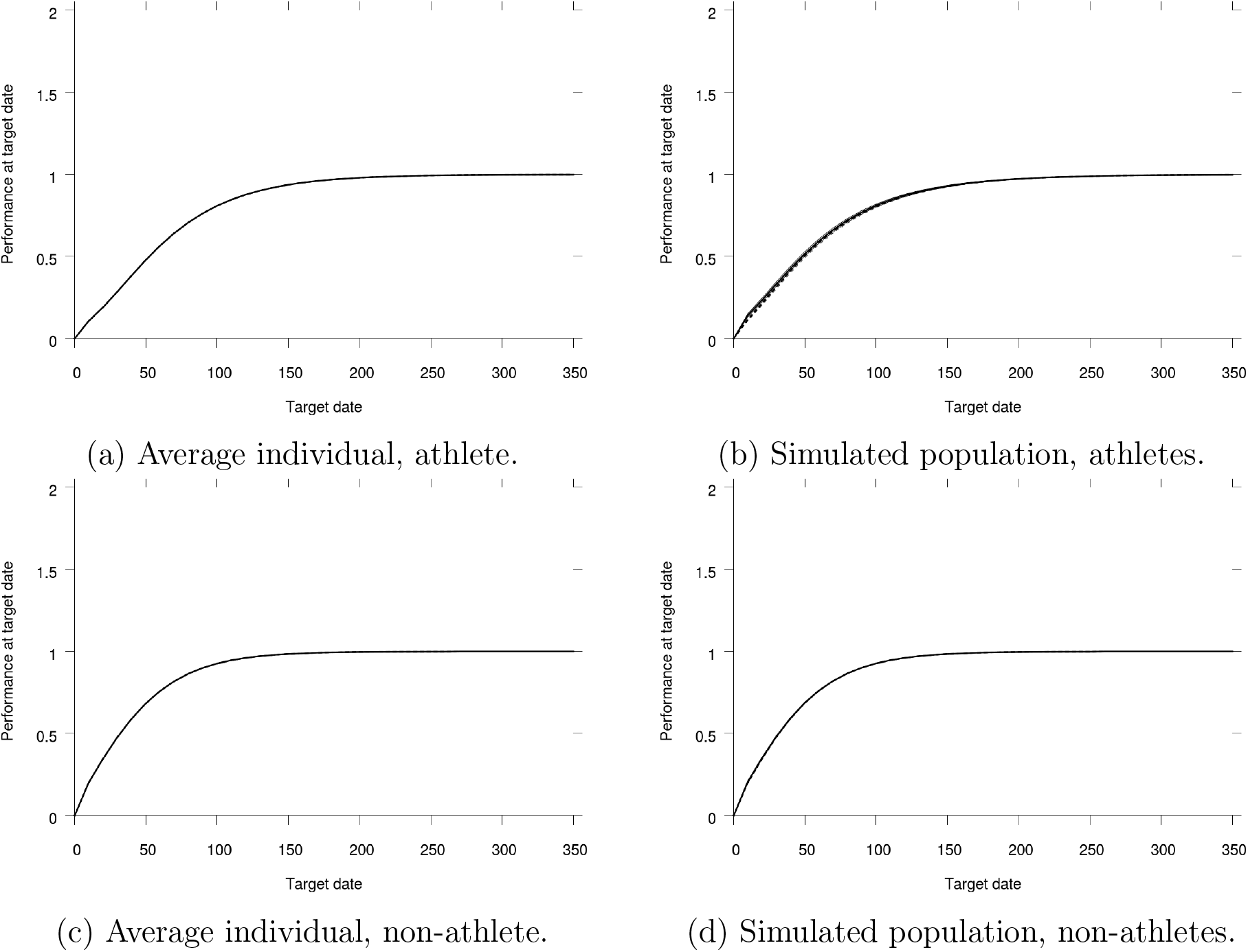
Performance of the OTDshare training strategy as a function of the target date *n* for the average athlete or non-athlete or for our simulated populations of 480 athletes and non-athletes. The dashed black line shows the expected performance of the best OTDshare training program when the individual parameter values are unknown. The plain black line shows the expected performance of the best OTDshare training programs when the individual parameter values are known. Light lines show the 95% confidence interval for our 480 simulated individuals.

**Figure App-2:**
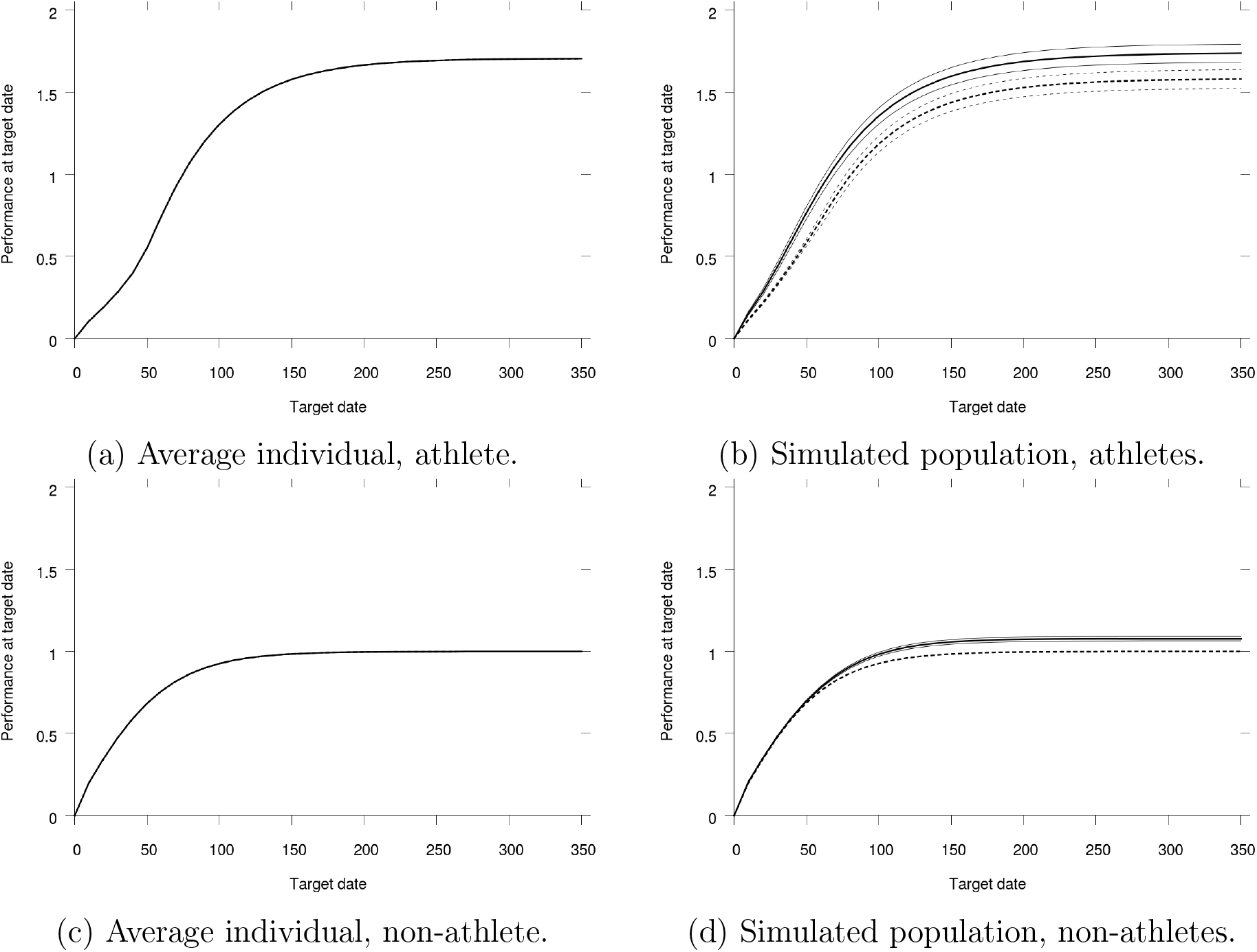
Performance of the Step training strategy as a function of the target date *n* for the average athlete or non-athlete or for our simulated populations of 480 athletes and non-athletes. The dashed black line shows the expected performance of the best Step training program when the individual parameter values are unknown. The plain black line shows the expected performance of the best Step training programs when the individual parameter values are known. Light lines show the 95% confidence interval for our 480 simulated individuals.

**Figure App-3:**
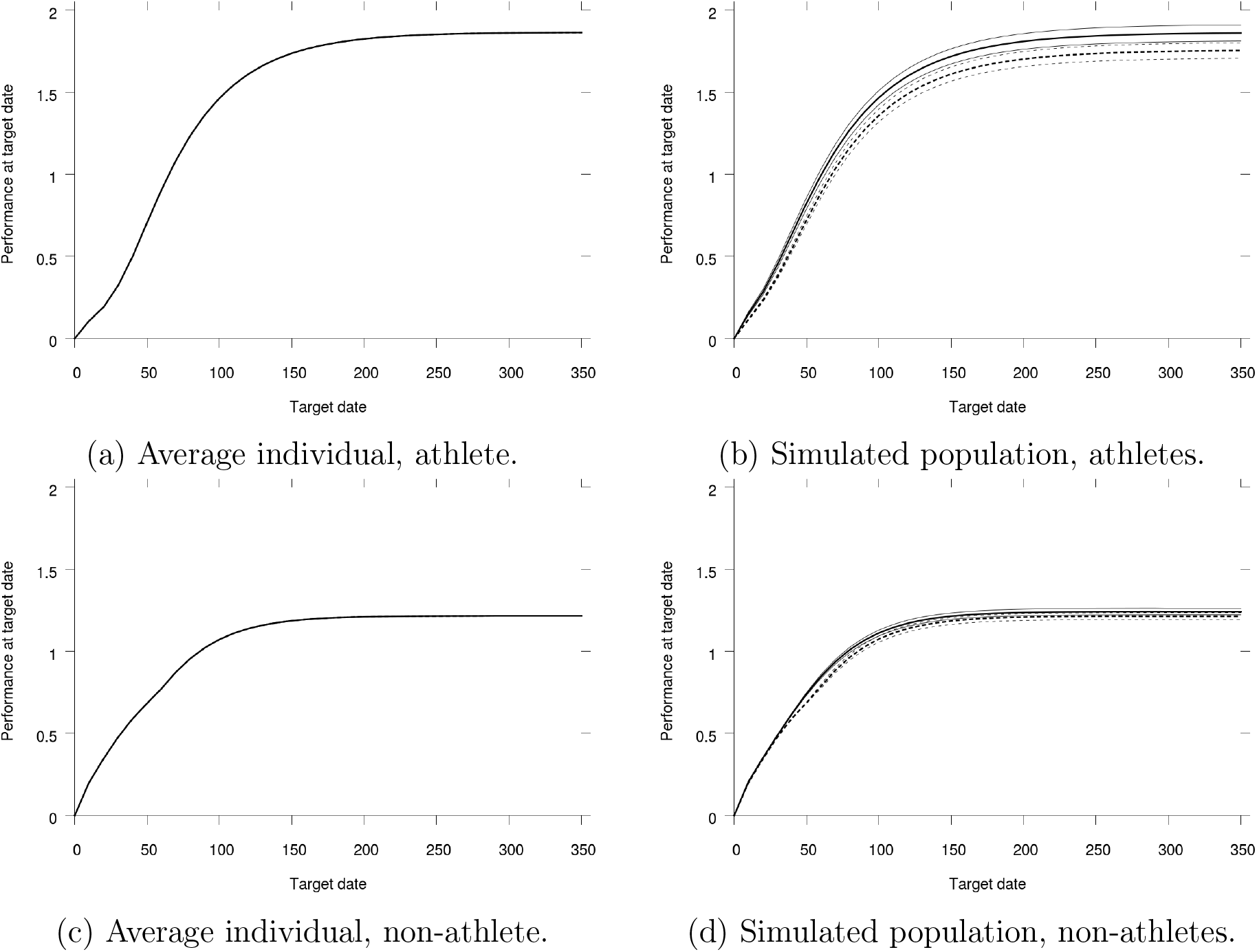
Performance of the Linear training strategy as a function of the target date *n* for the average athlete or non-athlete or for our simulated populations of 480 athletes and non-athletes. The dashed black line shows the expected performance of the best Linear training program when the individual parameter values are unknown. The plain black line shows the expected performance of the best Linear training programs when the individual parameter values are known. Light lines show the 95% confidence interval for our 480 simulated individuals.

**Figure App-4:**
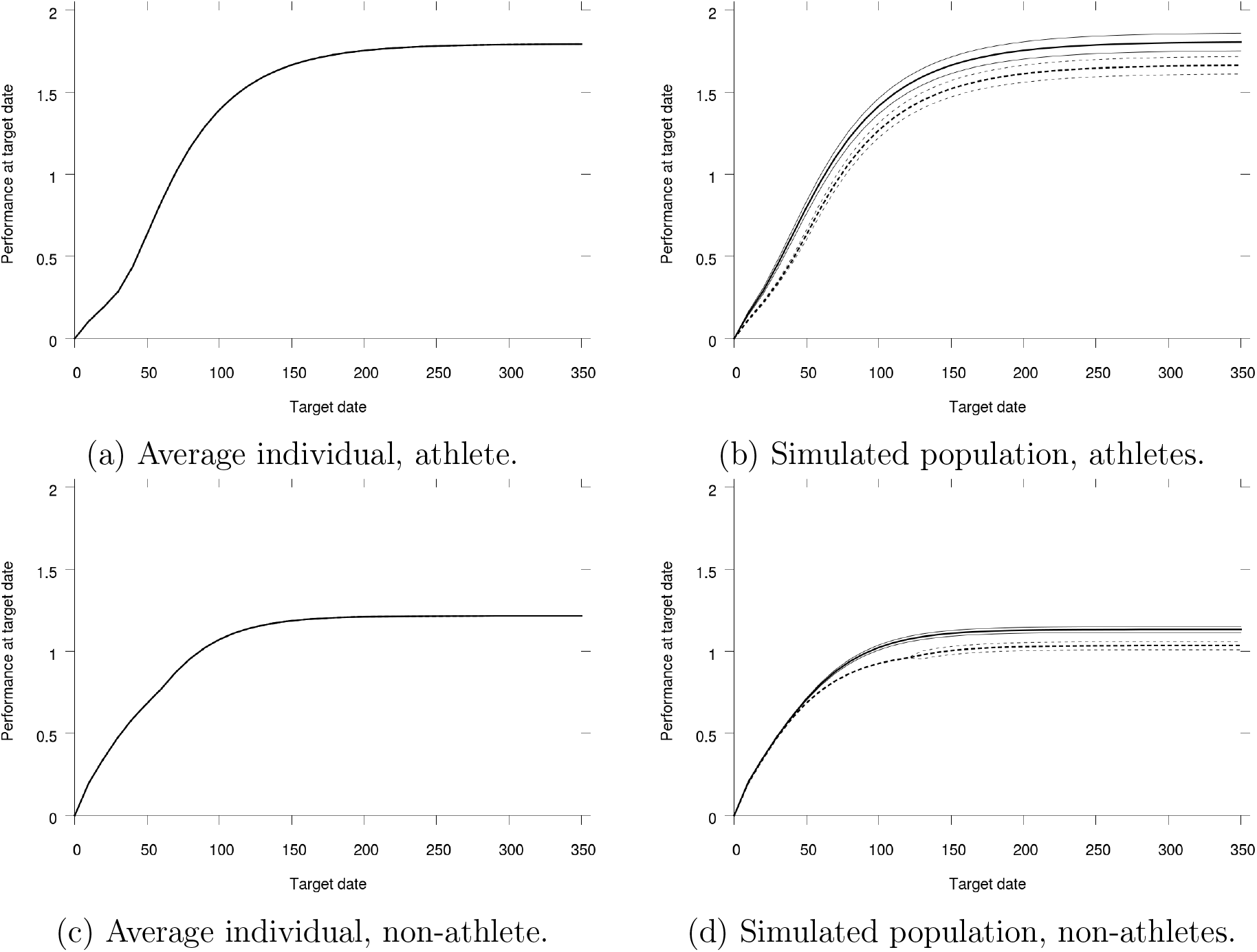
Performance of the Exponential training strategy as a function of the target date *n* for the average athlete or non-athlete or for our simulated populations of 480 athletes and non-athletes. The dashed black line shows the expected performance of the best Exponential training program when the individual parameter values are unknown. The plain black line shows the expected performance of the best Exponential training programs when the individual parameter values are known. Light lines show the 95% confidence interval for our 480 simulated individuals.

**Figure App-5:**
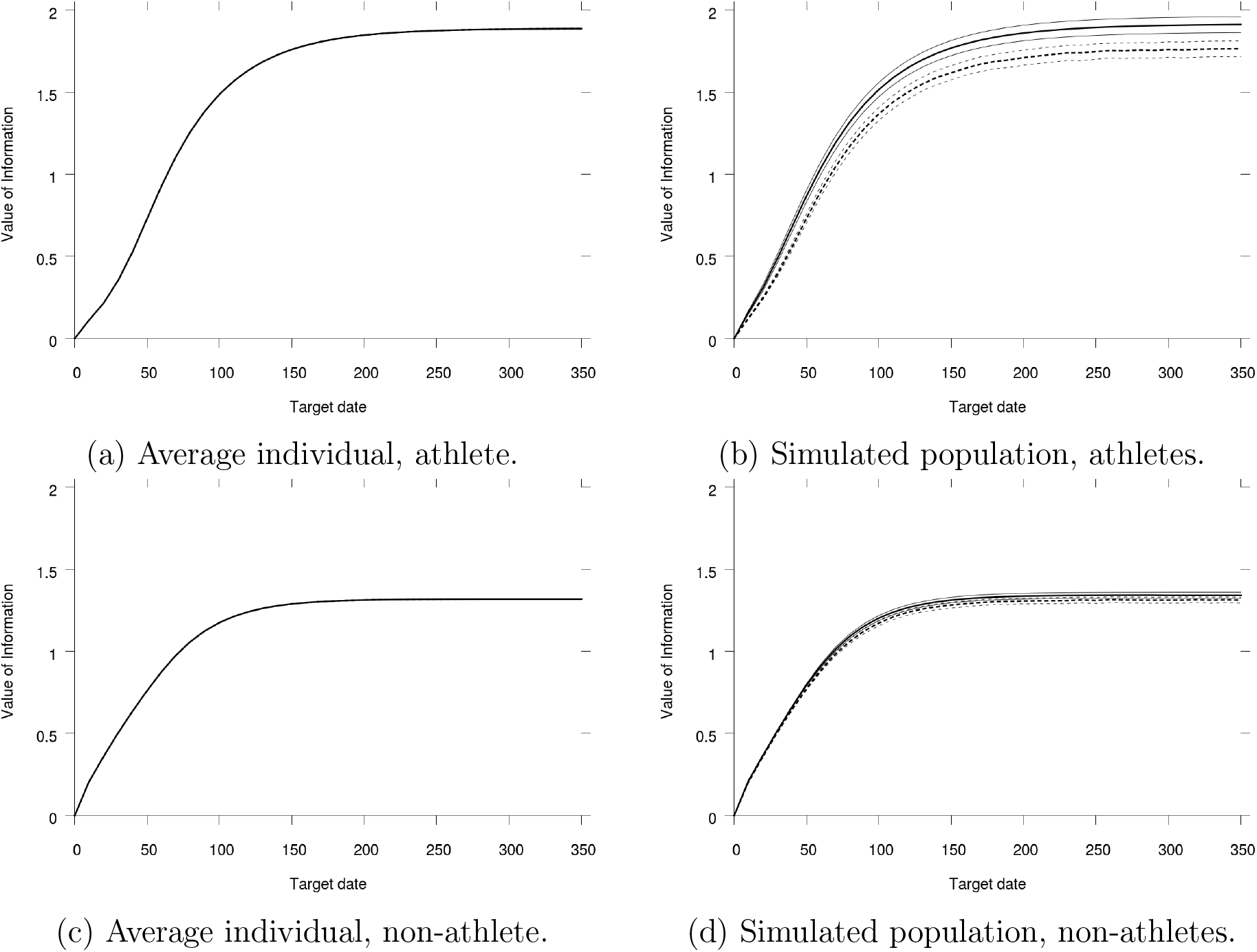
Performance of the MCTS training strategy as a function of the target date *n* for the average athlete or non-athlete or for our simulated populations of 480 athletes and non-athletes. The dashed black line shows the expected performance of the best MCTS training program when the individual parameter values are unknown. The plain black line shows the expected performance of the best MCTS training programs when the individual parameter values are known. Light lines show the 95% confidence interval for our 480 simulated individuals.

#### B.2 Best performances arguments

In Figures App-6, App-7, App-8 and App-8, we show the best arguments for the training strategies studied in this article.

**Figure App-6:**
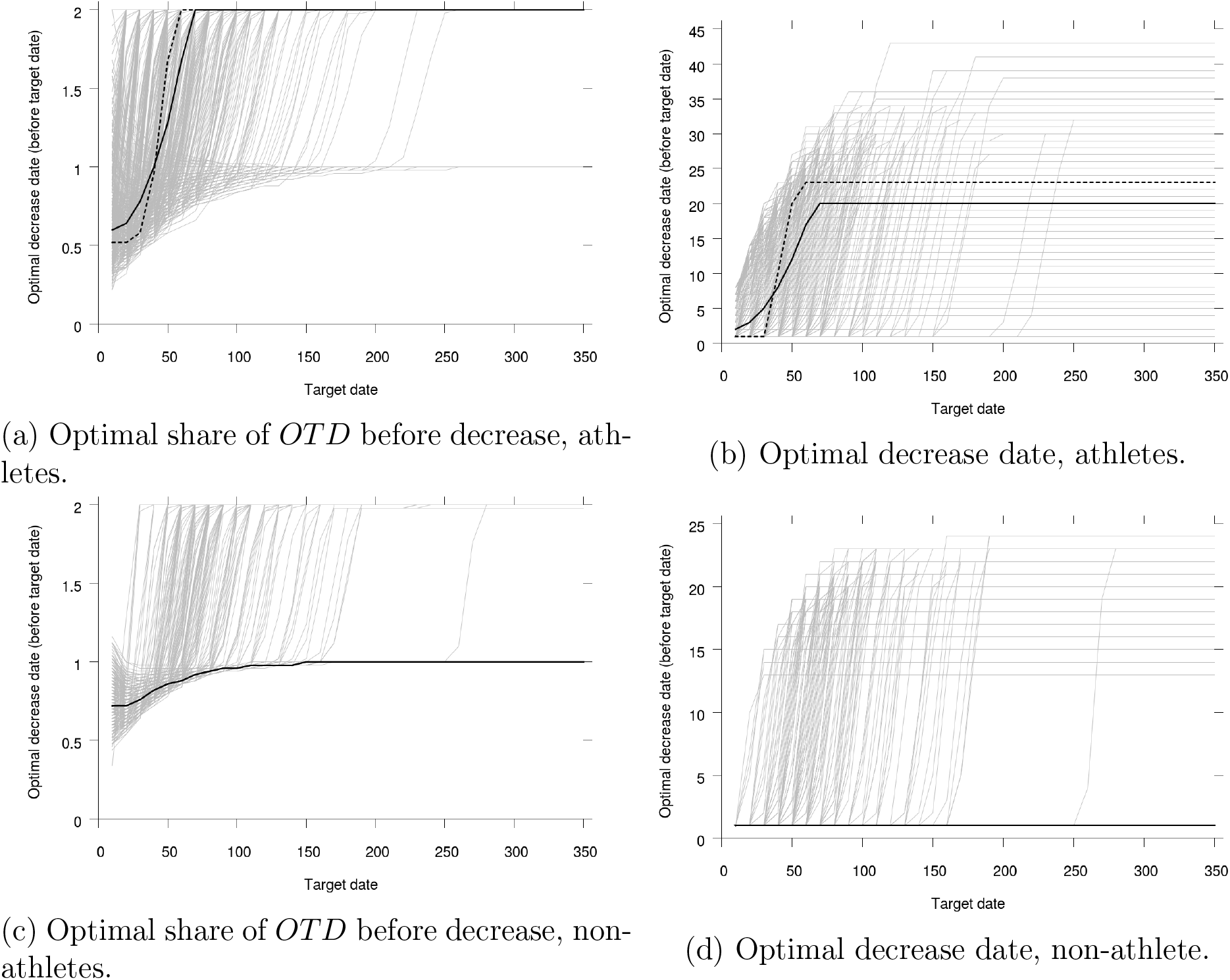
Share of *OTD* before decrease and decrease date – as a difference to the target date – (*q* and *n*′ with the notation of the main text) for the optimal Step training programs as functions of the target date *n* for athletes and non-athletes. Light lines for each of the 480 simulated individuals in the populations. Plain black line for the optimal Step training program for the populations without knowledge of individual parameter values. Dashed black line for the optimal Step training program for the average individual.

**Figure App-7:**
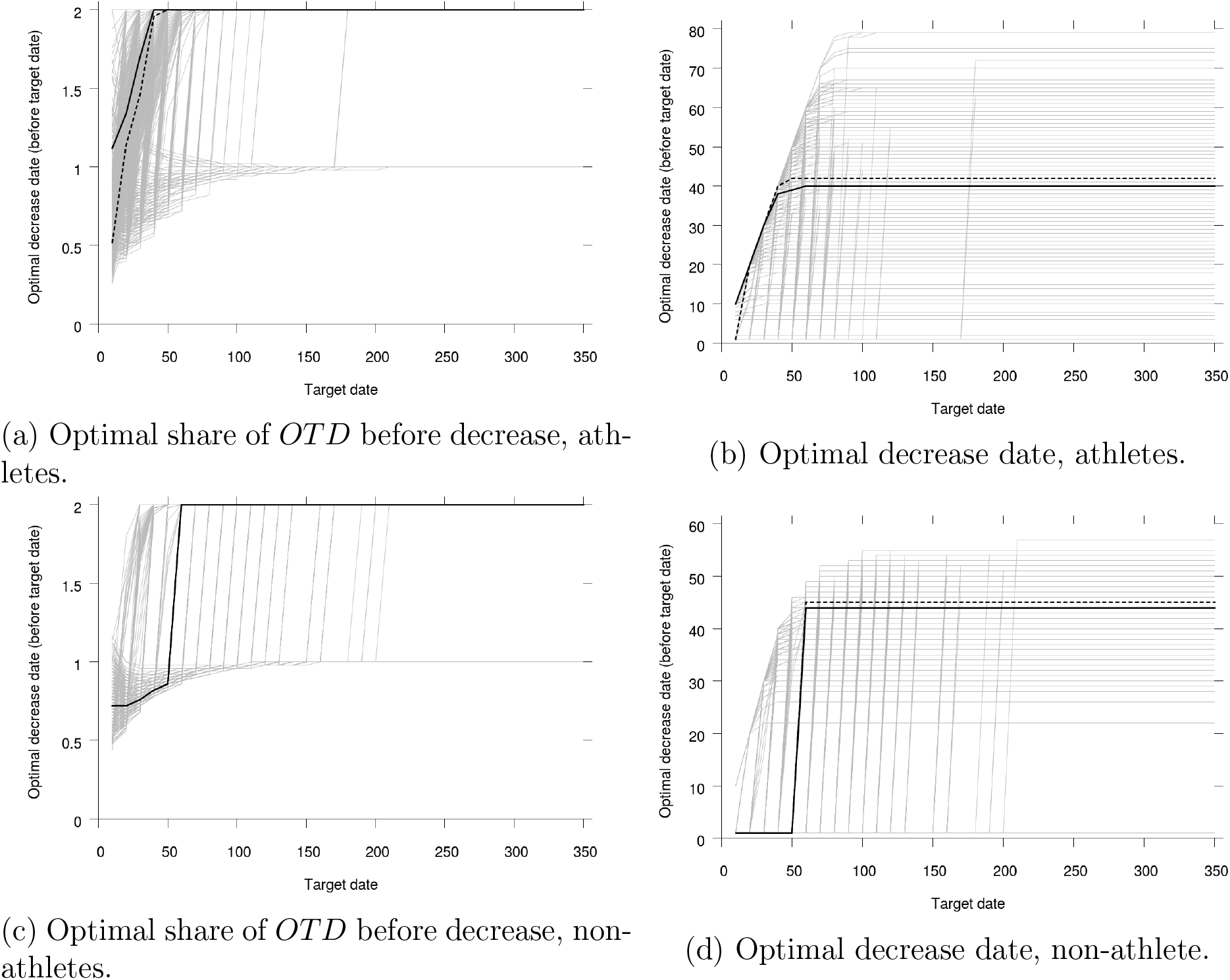
Share of *OTD* before decrease and decrease date – as a difference to the target date – (*q* and *n* −*n*′ with the notation of the main text) for the optimal Linear training program as functions of the target date *n* for athletes and non-athletes. Light lines for each of the 480 simulated individuals in the populations. Plain black line for the optimal Linear training program for the populations without knowledge of individual parameter values. Dashed black line for the optimal Linear training program for the average individual.

**Figure App-8:**
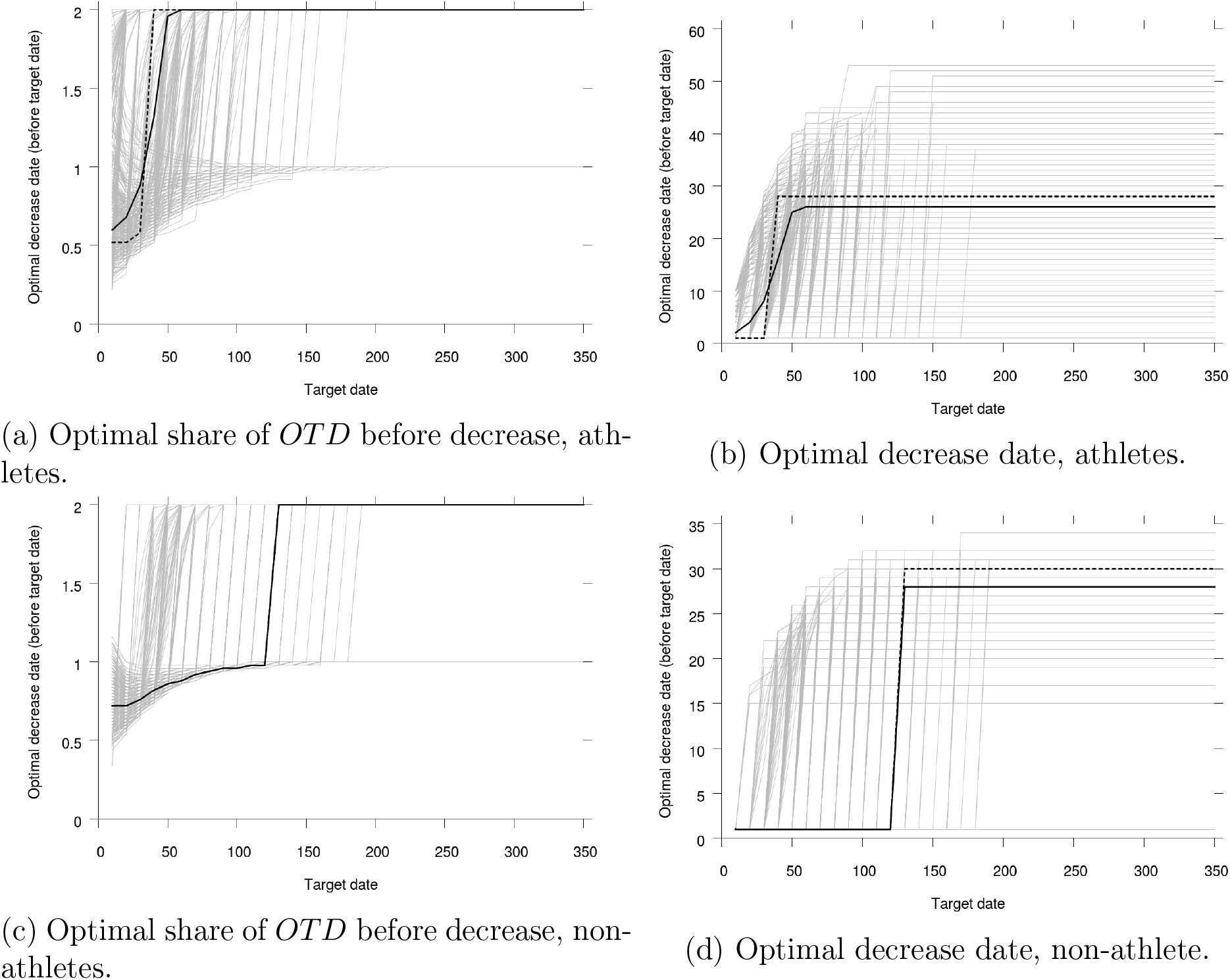
Share of *OTD* before decrease and decrease date – as a difference to the target date – (*q* and *n* −*n*′ with the notation of the main text) for the optimal Exponential training program as functions of the target date *n* for athletes and non-athletes. Light lines for each of the 480 simulated individuals in the populations. Plain black line for the optimal Exponential training program for the populations without knowledge of individual parameter values. Dashed black line for the optimal Exponential training program for the average individual.

**Figure App-9:**
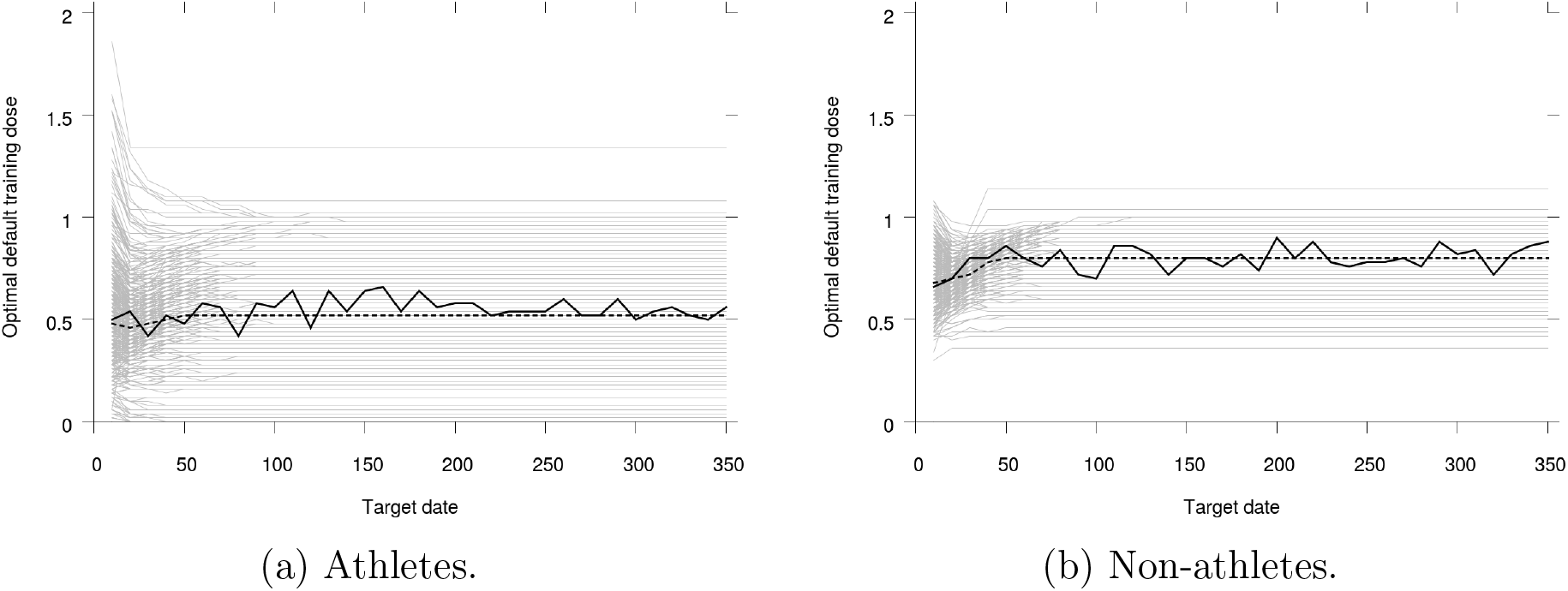
Default training dose for the optimal MCTS training programs as a function of the target date *n* for athletes and non-athletes. Light lines for each of the 480 simulated individuals in the populations. Plain black line for the optimal MCTS training program for the populations without knowledge of individual parameter values. Dashed black line for the optimal MCTS training program for the average individual.

#### B.3 Performances as functions of time

In Figures App-10, App-11, App-12, App-13 and App-14, we show the time profile of performance when the optimal training programs for the different strategies are implemented.

**Figure App-10:**
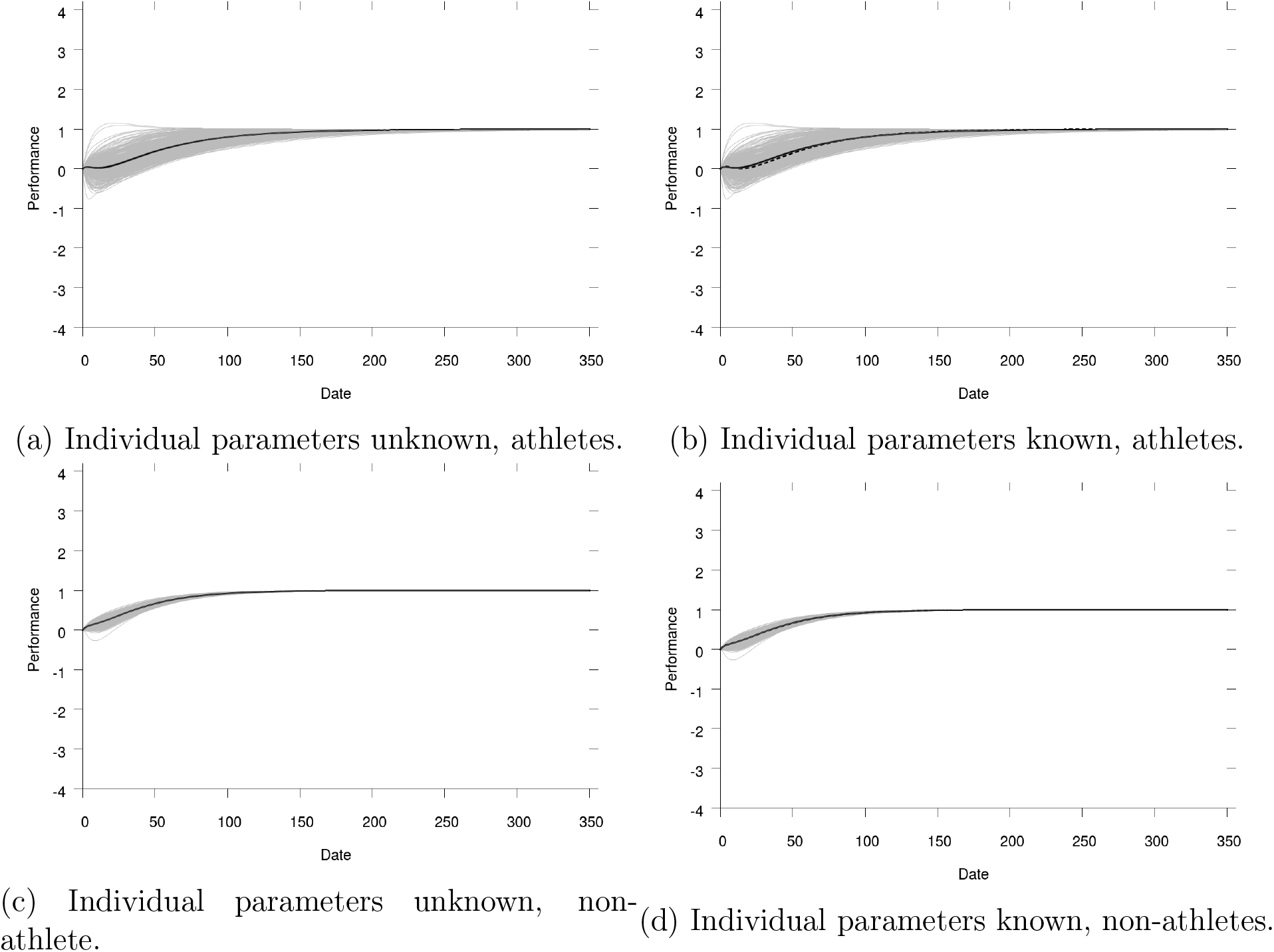
Performance depending on date for the OTDshare strategy for athletes and non-athletes with known or unknown individual parameters, target date 350. Very light lines for each of the individuals in the simulated population. Dashed line for average individual (only for known individual parameters), dark plain line for average performance across population. Light lines show the 95% confidence interval for our 480 simulated individuals.

**Figure App-11:**
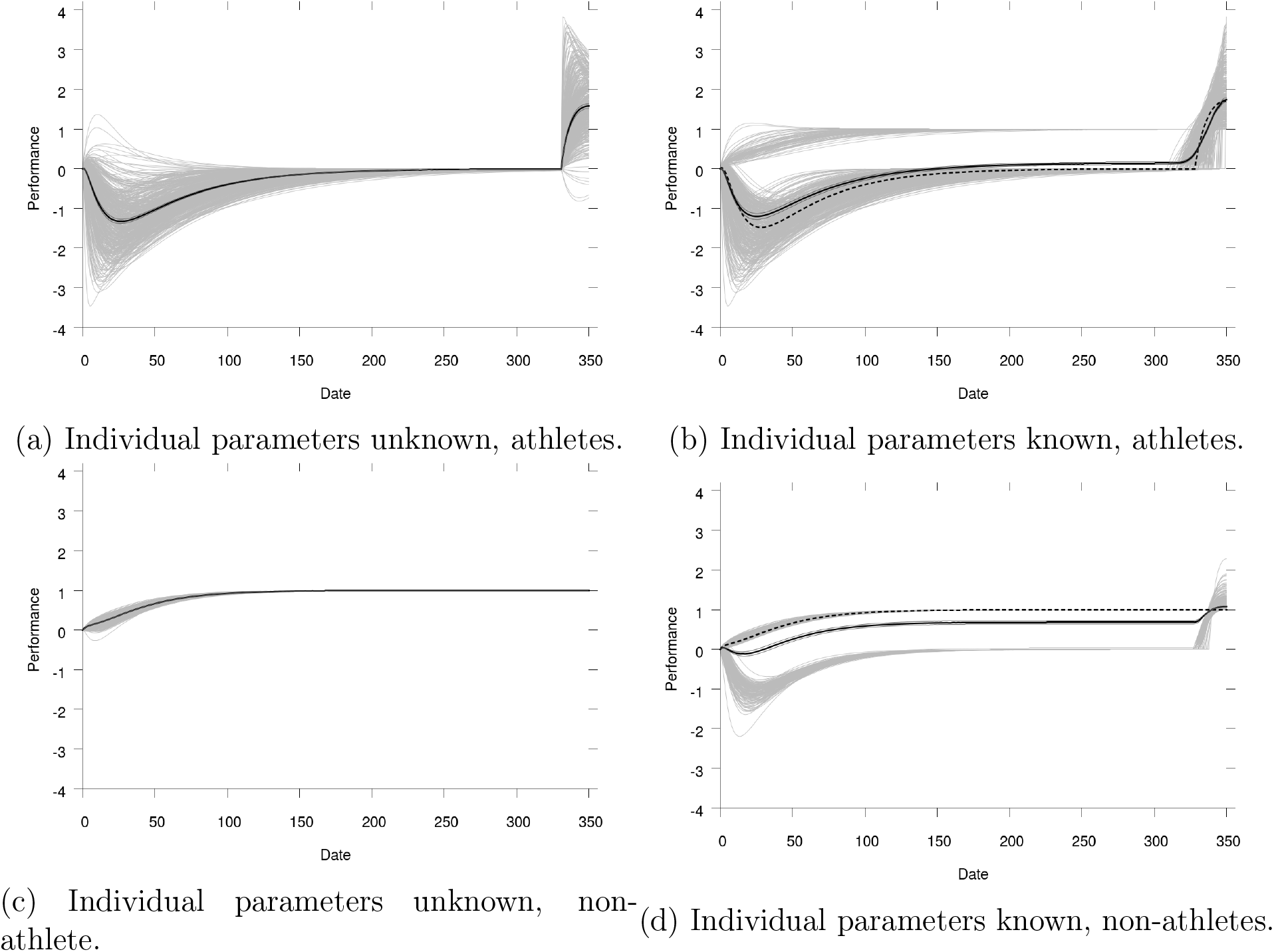
Performance depending on date for the Step strategy for athletes and non-athletes with known or unknown individual parameters, target date 350. Very light lines for each of the individuals in the simulated population. Dashed line for average individual (only for known individual parameters), dark plain line for average performance across population. Light lines show the 95% confidence interval for our 480 simulated individuals.

**Figure App-12:**
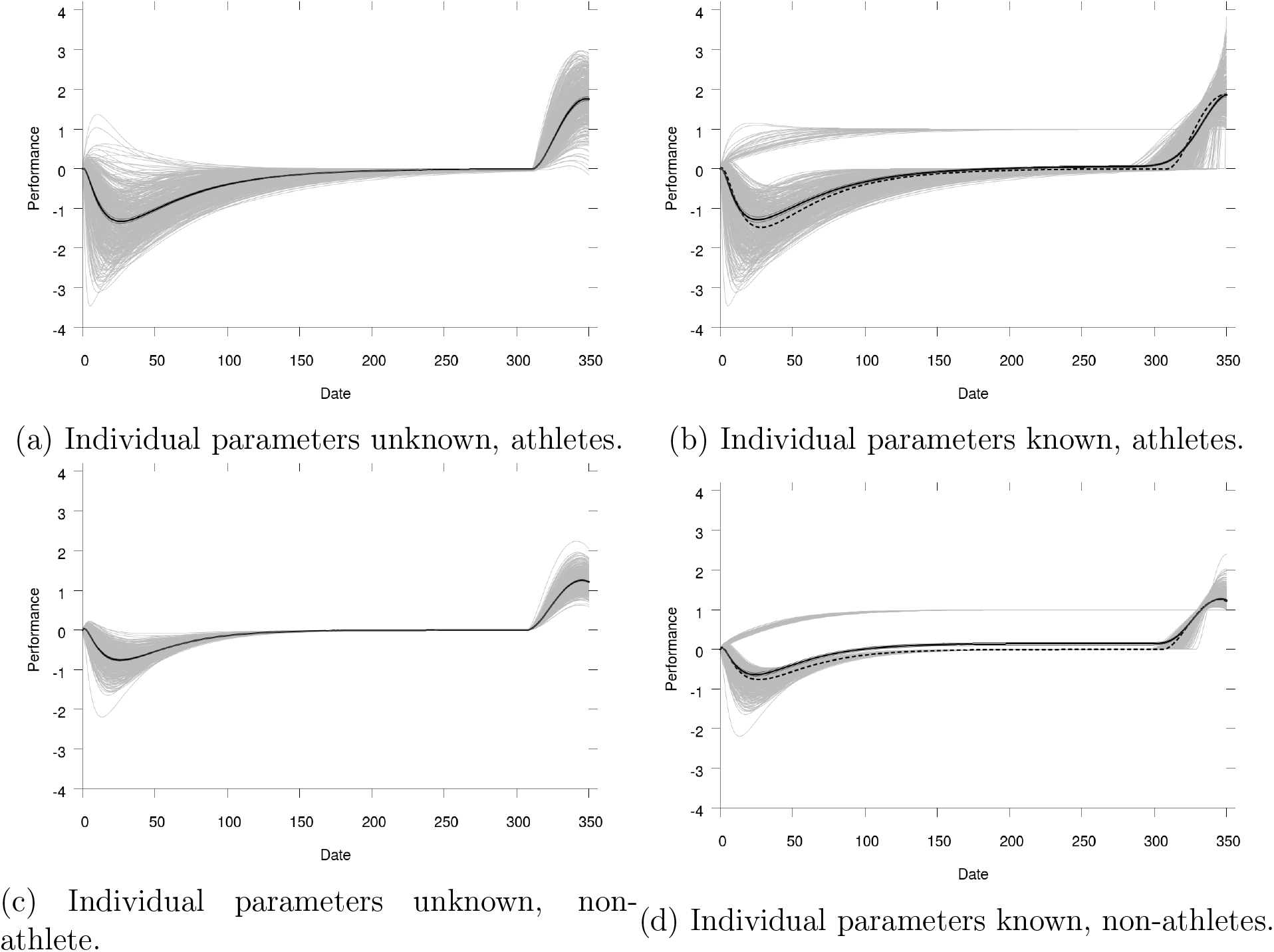
Performance depending on date for the Linear strategy for athletes and non-athletes with known or unknown individual parameters, target date 350. Very light lines for each of the individuals in the simulated population. Dashed line for average individual (only for known individual parameters), dark plain line for average performance across population. Light lines show the 95% confidence interval for our 480 simulated individuals.

**Figure App-13:**
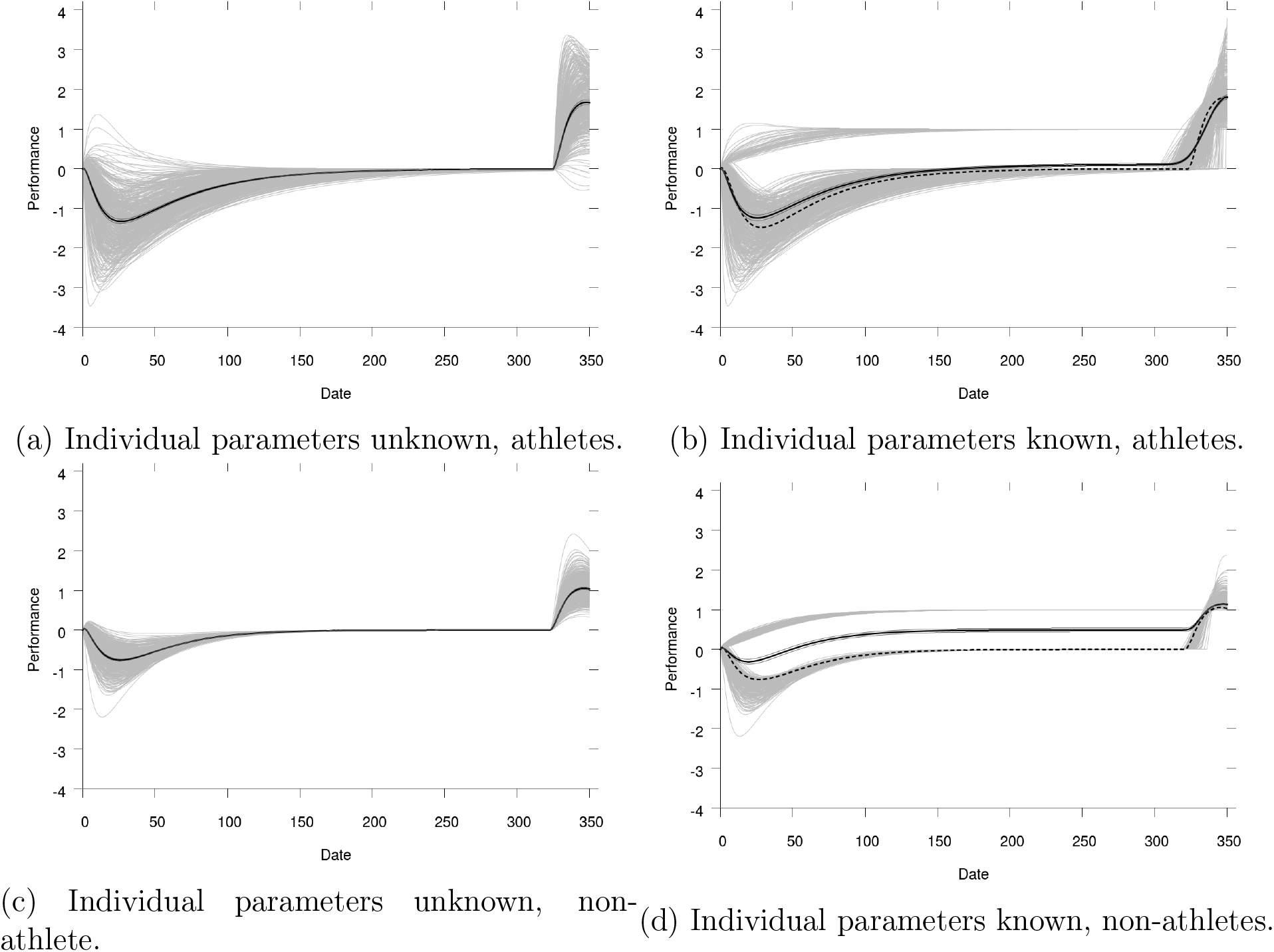
Performance depending on date for the Exponential strategy for athletes and non-athletes with known or unknown individual parameters, target date 350. Very light lines for each of the individuals in the simulated population. Dashed line for average individual (only for known individual parameters), dark plain line for average performance across population. Light lines show the 95% confidence interval for our 480 simulated individuals.

**Figure App-14:**
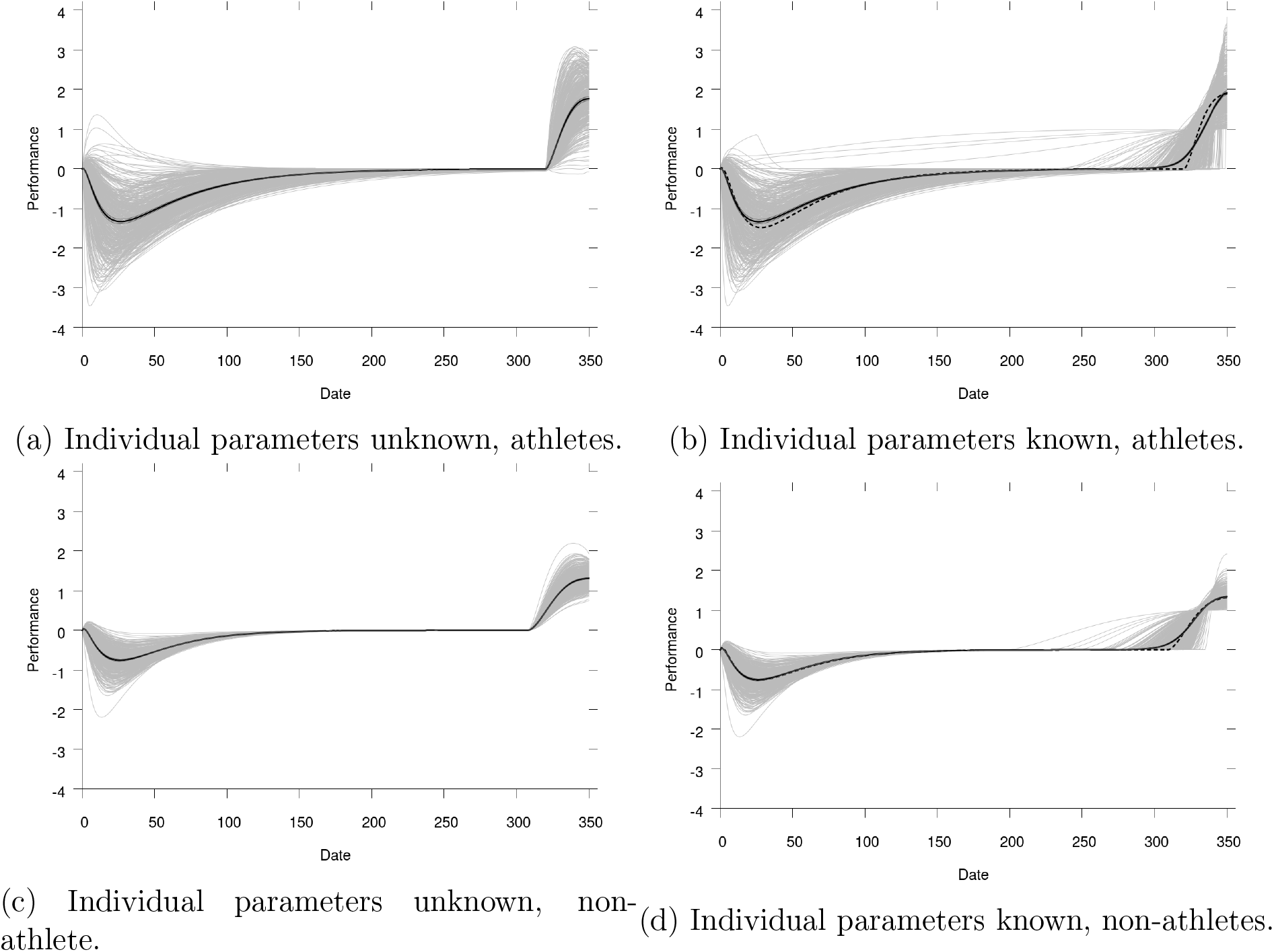
Performance depending on date for the MCTS strategy for athletes and non-athletes with known or unknown individual parameters, target date 350. Very light lines for each of the individuals in the simulated population. Dashed line for average individual (only for known individual parameters), dark plain line for average performance across population. Light lines show the 95% confidence interval for our 480 simulated individuals.

### C Differences in performance between MCTS and the other training strategies

In Figures App-15, App-17 and App-16, we show the difference in performance between the best MCTS training program and the best of each of the other training strategies studied in this article as functions of the target date.

**Figure App-15:**
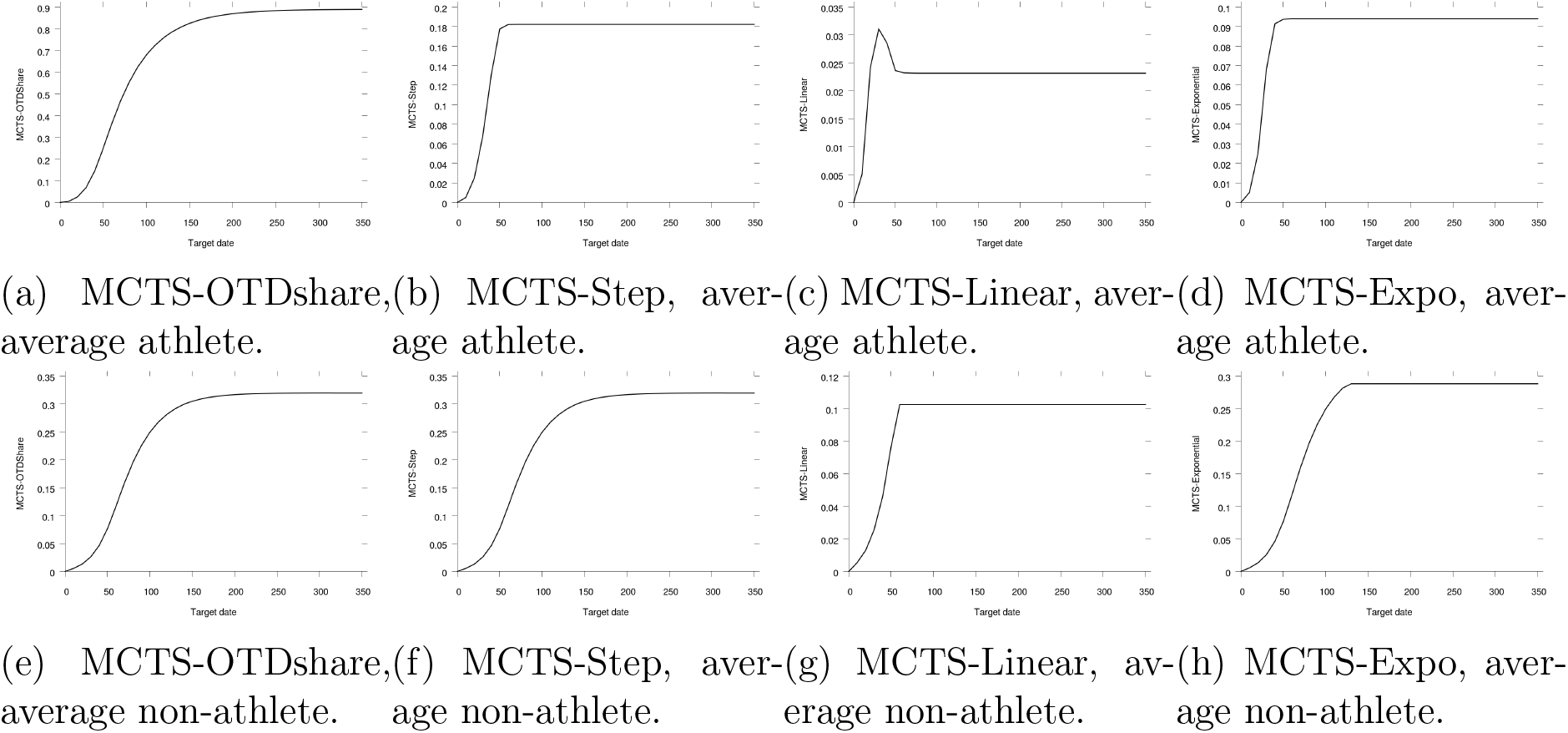
Difference in performance between the best MCTS training program and the best training programs for other strategies as functions of the training date for an average athlete and an average non-athlete. Light lines show the 95% confidence interval for our 480 simulated individuals.

**Figure App-16:**
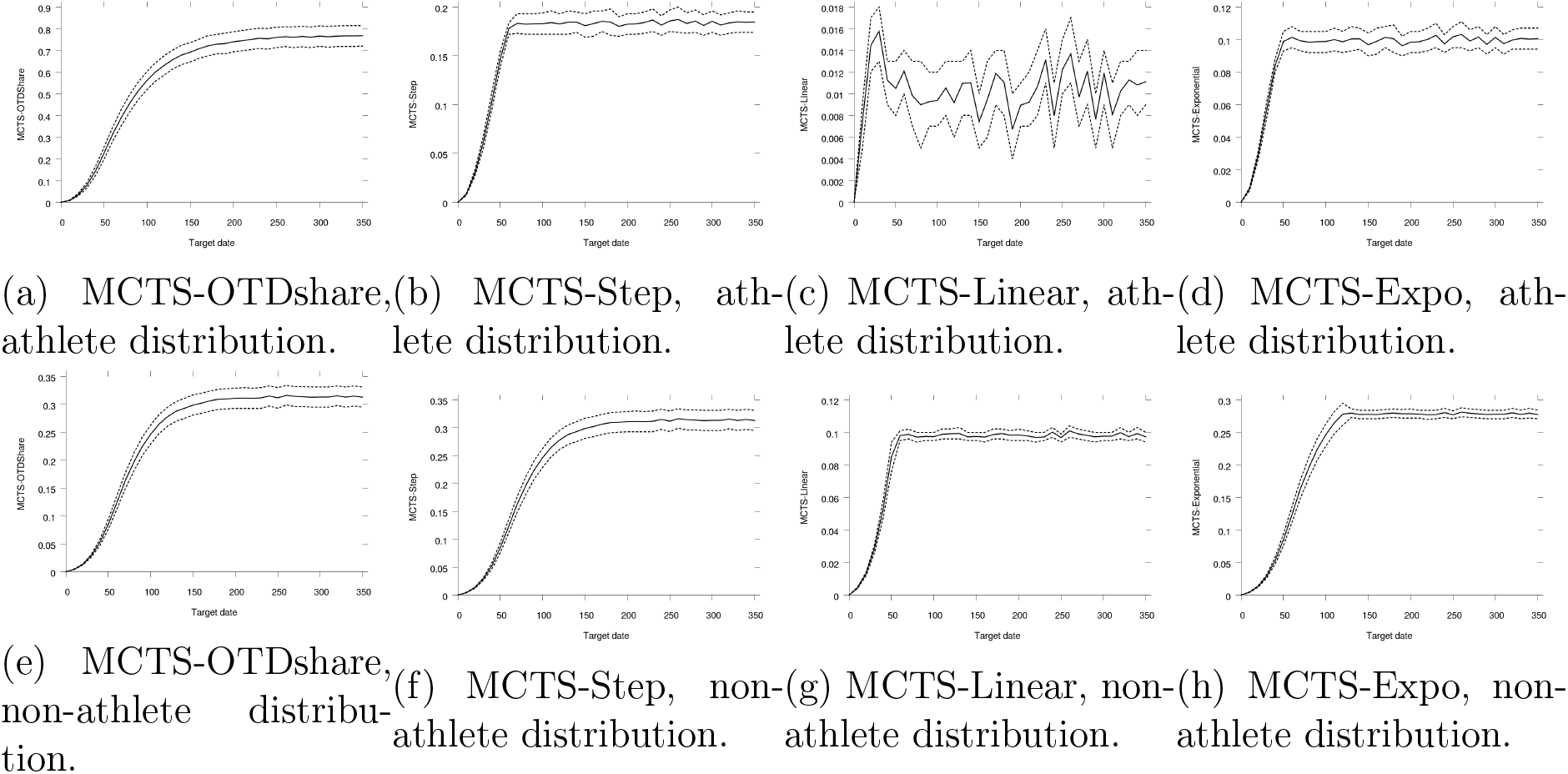
Difference in performance between the best MCTS training program and the best training programs for other strategies as functions of the training date for our simulated population of athletes and non-athletes with unknown individual parameter values. Light lines show the 95% confidence interval for our 480 simulated individuals.

**Figure App-17:**
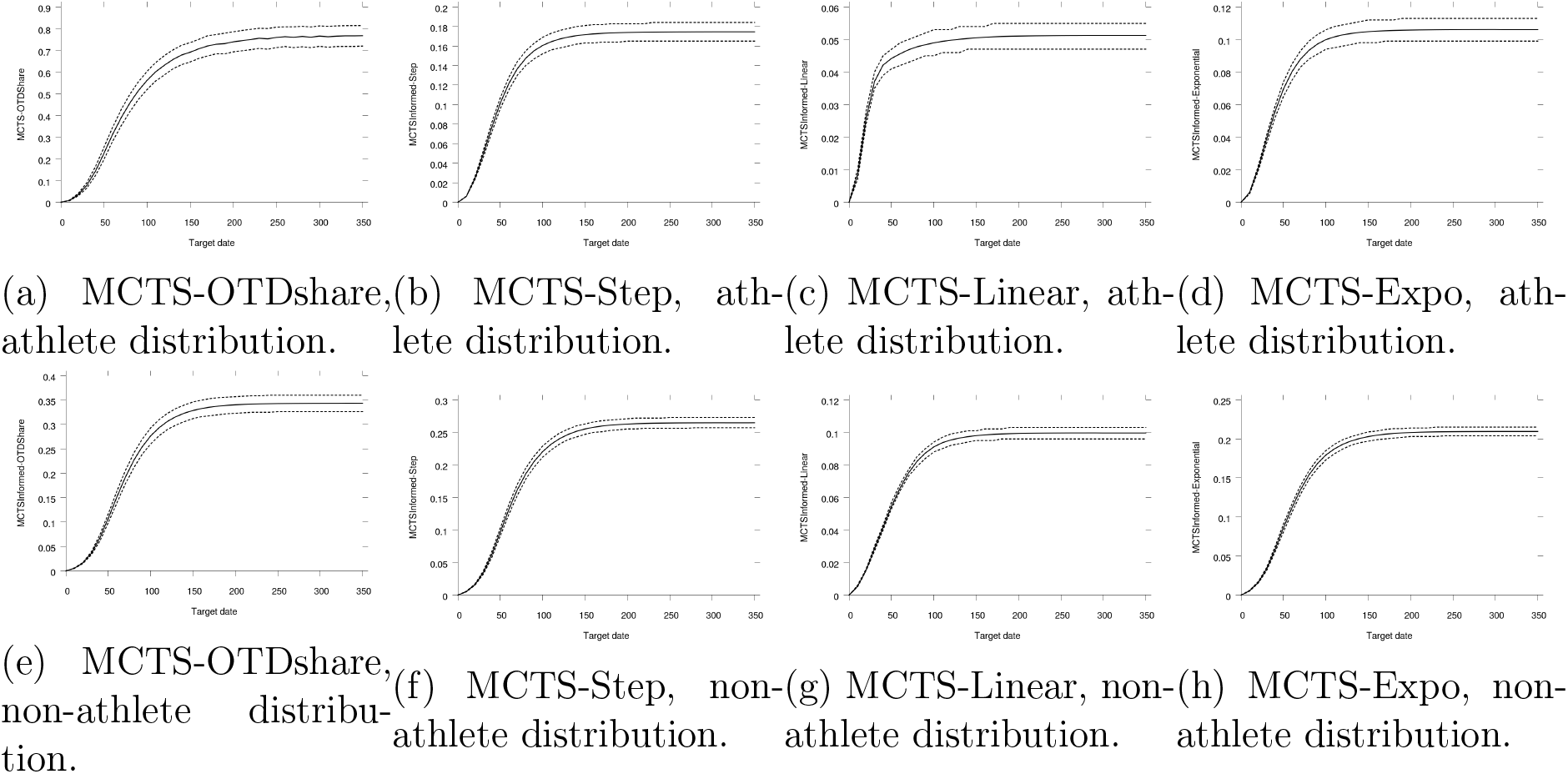
Difference in performance between the best MCTS training program and the best training programs for other strategies as functions of the training date for our simulated population of athletes and non-athletes with known individual parameter values. Light lines show the 95% confidence interval for our 480 simulated individuals.

### D Extension to gymnasts, skaters and swimmers

In this section, we extend the main results of our article to gymnasts, skaters and swimmers as specified in different articles in the literature, see Table App-1.

**Table App-1:** Parameter distributions for populations of athletes, non-athletes, gymnasts, skaters, swimmers.

| Parameter | Athletes<br>[27, 29] | Non-athletes<br>[4, 27, 29] | Gymnasts<br>[23] |
| --- | --- | --- | --- |
| $k_1$ | $\mathcal{N}(3.42 \times 10^{-2}, 9.6 \times 10^{-3})$ | $\mathcal{N}(3.1 \times 10^{-2}, 7.3 \times 10^{-3})$ | $\mathcal{N}(4.3 \times 10^{-2}, 2 \times 10^{-4})$ |
| $k_3$ | $\mathcal{N}(3.4 \times 10^{-3}, 3.3 \times 10^{-3})$ | $\mathcal{N}(3.5 \times 10^{-5}, 10^{-5})$ | $\mathcal{N}(6.2 \times 10^{-6}, 1.5 \times 10^{-6})$ |
| $\tau_1$ | $\mathcal{N}(43.6, 12.1)$ | $\mathcal{N}(30.8, 1.6)$ | $\mathcal{N}(60.4, 0.7)$ |
| $\tau_2$ | $\mathcal{N}(12.7, 7.2)$ | $\mathcal{N}(16.8, 3.3)$ | $\mathcal{N}(19.8, 0.46)$ |
| $\tau_3$ | $\mathcal{N}(4.1, 1.6)$ | $\mathcal{N}(2.3, 1)$ | $\mathcal{N}(5.5, 0.8)$ |
| Parameter | Skaters<br>[20] | Swimmers<br>[28] |  |
| $k_1$ | $\mathcal{N}(1.27 \times 10^{-6}, 3.36 \times 10^{-7})$ | $\mathcal{N}(3.15 \times 10^{-2}, 1.17 \times 10^{-2})$ | |
| $k_3$ | $\mathcal{N}(2.89 \times 10^{-10}, 1.24 \times 10^{-10})$ | $\mathcal{N}(4 \times 10^{-3}, 3.5 \times 10^{-3})$ | |
| $\tau_1$ | $\mathcal{N}(39.9, 6.8)$ | $\mathcal{N}(45.6, 12.7)$ | |
| $\tau_2$ | $\mathcal{N}(5.2, 1.1)$ | $\mathcal{N}(11.3, 7.9)$ | |
| $\tau_3$ | $\mathcal{N}(1.9, 1.2)$ | $\mathcal{N}(3.8, 1.8)$ | |

**Table App-2:**
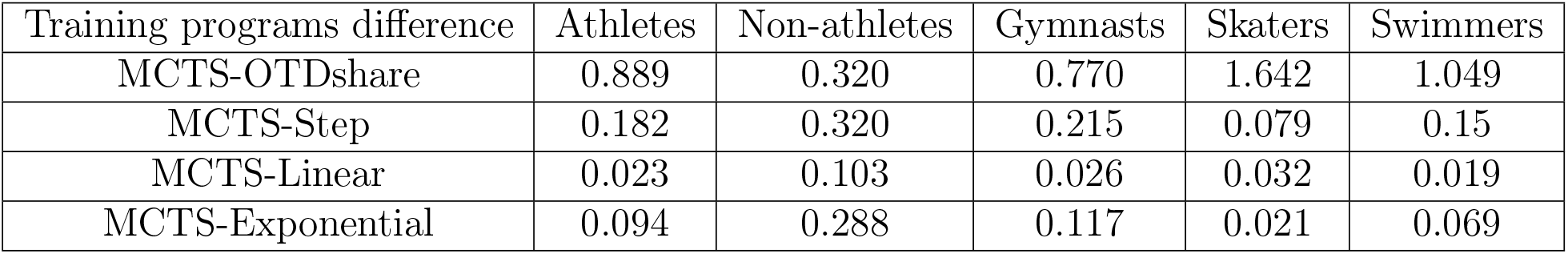
Difference in performance between the best MCTS training program and the best other training programs for an average athletes, average non-athletes, average gymnasts, average skaters, average swimmers and for a target date of 350.

**Table App-3:**
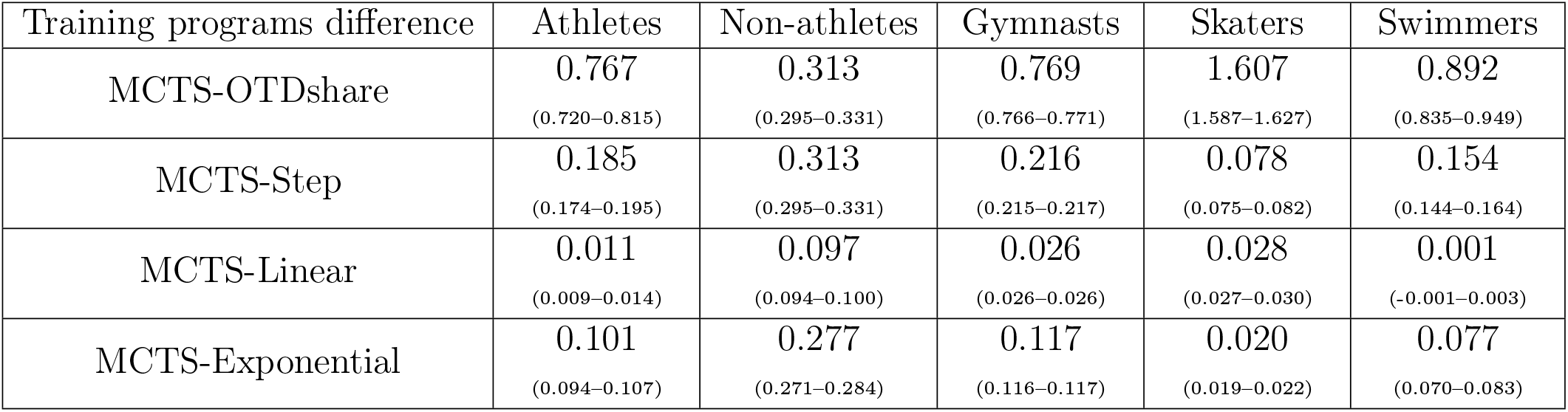
Difference in performance between the best MCTS training program and the best other training programs for athletes, non-athletes, gymnasts, skaters, swimmers and for a target date of 350. Individual parameter values are unknown. 95% confidence interval for our 480 simulated individuals in parenthesis.

| Training programs difference | Athletes | Non-athletes | Gymnasts | Skaters | Swimmers |
| --- | --- | --- | --- | --- | --- |
| MCTS-OTDshare | **0.767**<br>(0.720-0.815) | **0.313**<br>(0.295-0.331) | **0.769**<br>(0.766-0.771) | **1.607**<br>(1.587-1.627) | **0.892**<br>(0.835-0.949) |
| MCTS-Step | **0.185**<br>(0.174-0.195) | **0.313**<br>(0.295-0.331) | **0.216**<br>(0.215-0.217) | **0.078**<br>(0.075-0.082) | **0.154**<br>(0.144-0.164) |
| MCTS-Linear | **0.011**<br>(0.009-0.014) | **0.097**<br>(0.094-0.100) | **0.026**<br>(0.026-0.026) | **0.028**<br>(0.027-0.030) | **0.001**<br>(-0.001-0.003) |
| MCTS-Exponential | **0.101**<br>(0.094-0.107) | **0.277**<br>(0.271-0.284) | **0.117**<br>(0.116-0.117) | **0.020**<br>(0.019-0.022) | **0.077**<br>(0.070-0.083) |

**Table App-4:**
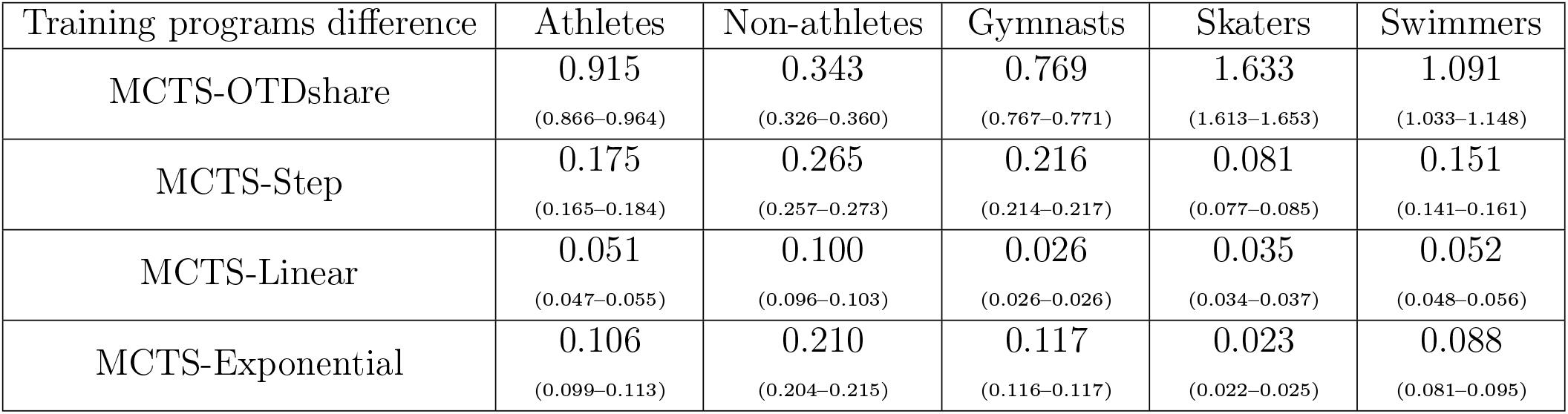
Difference in performance between the best MCTS training program and the best other training programs for athletes, non-athletes, gymnasts, skaters, swimmers and for a target date of 350. Individual parameter values are known. 95% confidence interval for our 480 simulated individuals in parenthesis.

| Training programs difference | Athletes | Non-athletes | Gymnasts | Skaters | Swimmers |
| --- | --- | --- | --- | --- | --- |
| MCTS-OTDshare | **0.915**<br>(0.866-0.964) | **0.343**<br>(0.326-0.360) | **0.769**<br>(0.767-0.771) | **1.633**<br>(1.613-1.653) | **1.091**<br>(1.033-1.148) |
| MCTS-Step | **0.175**<br>(0.165-0.184) | **0.265**<br>(0.257-0.273) | **0.216**<br>(0.214-0.217) | **0.081**<br>(0.077-0.085) | **0.151**<br>(0.141-0.161) |
| MCTS-Linear | **0.051**<br>(0.047-0.055) | **0.100**<br>(0.096-0.103) | **0.026**<br>(0.026-0.026) | **0.035**<br>(0.034-0.037) | **0.052**<br>(0.048-0.056) |
| MCTS-Exponential | **0.106**<br>(0.099-0.113) | **0.210**<br>(0.204-0.215) | **0.117**<br>(0.116-0.117) | **0.023**<br>(0.022-0.025) | **0.088**<br>(0.081-0.095) |

**Table App-5:** Value of information for different training strategies at target date 350. 95% confidence interval for our 480 simulated individuals in parenthesis.

| Training strategy | Athletes | Non-athletes | Gymnasts | Skaters | Swimmers |
| --- | --- | --- | --- | --- | --- |
| OTDshare | 0.000<br>(0.000-0.000) | 0.000<br>(0.000-0.000) | 0.000<br>(0.000-0.000) | 0.000<br>(0.000-0.000) | 0.000<br>(0.000-0.000) |
| Step | 0.157<br>(0.134-0.181) | 0.078<br>(0.064-0.093) | 0.000<br>(0.000-0.000) | 0.023<br>(0.020-0.027) | 0.201<br>(0.175-0.227) |
| Linear | 0.107<br>(0.090-0.124) | 0.028<br>(0.023-0.033) | 0.000<br>(0.000-0.000) | 0.019<br>(0.016-0.021) | 0.147<br>(0.127-0.167) |
| Exponential | 0.142<br>(0.120-0.164) | 0.098<br>(0.087-0.110) | 0.000<br>(0.000-0.000) | 0.023<br>(0.020-0.026) | 0.187<br>(0.163-0.210) |
| MCTS | 0.147<br>(0.128-0.167) | 0.030<br>(0.026-0.034) | 0.000<br>(0.000-0.000) | 0.026<br>(0.023-0.029) | 0.198<br>(0.176-0.221) |

## Footnotes

1 Some physiological correlates have been found *ex-post*, see [2, 7, 32] for instances.

2 Results are given in percentage points of the performance obtained with stationary optimal training dose (pp POTD).

3 In all this article, we will compare performances obtained by different training programs. Hence, with no loss of generality, we will assume *p*° = 0.

4 We also computed results with the population parameter values distributions for gymnasts [23], skaters [20] and swimmers [28]. Results are given in Appendix.

5 Let us clarify a terminology point here. We will call a training program a set of impulses. We will call a training strategy a family of training programs. For instance, the OTDshare(1,350) training program belongs to the OTDshare training strategy.

6 Notice that, by definition, the Step(*q, n* −1, *n*) training program is the same as the OTDshare(*q,n*) training program.

7 Notice that, by definition, the Linear(*q, n* −1, *n*) training program is the same as the OTDshare(*q,n*) training program.

8 Notice that, by definition, the Exponential(*q, n* −1, *n*) training program is the same as the OTDshare(*q,n*) training program.

9 For the sake of simplicity, we omit the dependence on the individual parameter values.

10 Throughout, performance is expressed relatively to *POTD*.

11 In Appendix, we show similar figures for the Step training strategy (Figures App-2, App-6), the Linear training strategy (Figures App-3, App-7) and the Exponential training strategy (Figures App-4, App-8). The corresponding performance profiles for those strategies and the others studied in the remainder of this article are given in Figures App-10, App-11, App-12, App-13 and App-14.

12 Notice that the value of information is irrelevant when we consider only average individuals of which we necessarily know the parameter values.

13 Main results as functions of the target date are displayed in Appendix.

14 Notice that, by definition, Step(349,1,350) is precisely OTDshare(1,350). Hence the Step strategy offers no improvement over the OTDshare strategy in this case.

15 With no surprise but still performing a powerful test, this statement is actually true for all target dates (see Section C in Appendix), all informational contexts (see the remainder of this article) and also for gymnasts, skaters and swimmers (see Section D in Appendix).

16 As in [24], there is no constraint in our model, on this overload amount of work long before the target date.

17 The MCTS algorithm must be slightly modified in this case where the individual parameter values are unknown. These modifications are described in Section A in Appendix.

18 Hence, for non-athletes, the best Step training program is, again, an OTDshare training program.

19 Specifically, we draw a population of 480 individuals in all our simulations.

